# Migraine, Neurogenic Inflammation, and Cardiovascular Risk: A Pharmacoepidemiologic Study of ACEI/ARB and Anti-CGRP Treatment

**DOI:** 10.1101/2025.06.02.25328819

**Authors:** Kaicheng Wang, Brenda T. Fenton, John P. Ney, Vinh X. Dao, Haidong Lu, Alexander B. Guirguis, Emmanuelle A. Schindler, Adam de Havenon, Melissa Skanderson, Sarah E. Anthony, Laura J. Burrone, Jason J. Sico

## Abstract

**Background:** Migraine is a prevalent neurological condition associated with increased cardiovascular risk; however, the pathophysiologic mechanisms underlying this association remains poorly understood, and the long-term cardiovascular effects of migraine preventive medications have yet to be determined.

**Methods:** We emulated two separate two-arm target trials comparing patients who initiated lisinopril/candesartan or anti-calcitonin gene-related peptide (aCGRP) treatments versus those receiving topiramate for migraine prevention within the U.S. Department of Veterans Affairs between June 1, 2018 and September 30, 2024. The outcome was major adverse cardiovascular event (MACE) including myocardial infarction (MI), ischemic stroke, intracerebral and subarachnoid hemorrhage (ICH/SAH) and all-cause mortality. Five-year cumulative incidences, risk differences, risk ratios and overall hazard ratios (HRs) were estimated.

**Results:** Among 48,610 patients initiating lisinopril/candesartan (mean [SD] age, 52.3 [12.3] years; 18.9% women) and 25,635 initiating aCGRP treatment (mean [SD] age, 47.2 [12.2] years; 39.4% women), lisinopril/candesartan was associated with a higher risk of MACE (HR 1.21; 95%CI, 1.10-1.34), particularly MI (HR 1.28; 95%CI, 1.03-1.59). The MI risk was greater in patients without documented cardiovascular indications (HR 2.75; 95%CI, 2.01-3.77), patients aged <40 years (HR 3.18; 95%CI, 1.63-6.20), or with baseline systolic blood pressure <130 mm Hg (HR 1.86; 95%CI, 1.44-2.39). aCGRP use was associated with a reduced risk of MI (HR 0.80; 95% CI, 0.62-1.02) and was not associated with ischemic stroke (HR 1.11; 95% CI, 0.84-1.49) or ICH/SAH (HR 0.95; 95% CI, 0.65-1.38).

**Conclusions:** In this retrospective cohort study, lisinopril/candesartan may increase cardiovascular risk among migraine patients, particularly those at low risk of atherosclerosis, while aCGRP treatment appears safe and potentially protective against MI. These findings suggest a role for neurogenic inflammation–mediated by CGRP and substance P–in migraine-associated, non-atherosclerotic cardiovascular risk. Future research is warranted.

## Introduction

Migraine is a prevalent and incapacitating neurological disorder that affects 15.9% of adults in the United States.^1^ Epidemiologic studies^2^ have showed that individuals with migraine, particularly those with aura^3^ and frequent attacks^4^, are at increased risk of myocardial infarction (MI) and ischemic stroke. Although the underlying pathophysiology remains unclear, current state of knowledge supports a cascade in which cortical spreading depression activates the trigeminovascular system, leading to the release of neuropeptides such as calcitonin gene-related peptide (CGRP) and substance P (SP).^5, 6^ CGRP promotes vasodilation and proinflammatory responses, while SP contributes to pain transmission and neurogenic inflammation.^7, 8^ Together, these observations support the hypothesis that migraine-associated cardiovascular risk may be mediated, at least in part, by neurogenic inflammation triggered during migraine attacks.

Meanwhile, the susceptibility of cardiovascular events complicates the selection of effective migraine treatments. Several conventional abortive and preventive medications are contraindicated or must be used with caution in patients with cardiovascular risk. For example, triptan use is associated with at least a threefold short-term risk of major adverse cardiovascular events (MACE)^9^, and tricyclic antidepressants are associated with a 20% higher risk of MACE in patients with chronic pain^10^. Among novel preventive treatments, monoclonal antibodies and small-molecule antagonists that block CGRP or its receptor (aCGRP treatments) are now recommended as first-line treatment for migraine prevention.^11^ However, blocking CGRP could theoretically lead to vasoconstriction. Alternatively, lisinopril, an angiotensin-converting enzyme inhibitor (ACEI), and candesartan, an angiotensin II receptor blocker (ARB), are also guideline-recommended for migraine prevention,^12, 13^ though they remain underutilized without comorbid hypertension. Notably, ACE degrades SP; thus, ACE inhibition by lisinopril may reduce the breakdown of SP, potentially amplify neuroinflammatory responses. Despite this, ACEIs and ARBs may offer dual benefits for patients with migraine and cardiovascular risk, as meta-analysis have showed that they reduce cardiovascular risk and mortality by 11% among individuals with or at risk for atherosclerotic disease.^14^

Given the uncertainty surrounding the long-term cardiovascular effects of these drug classes in patients with migraine, we performed a pharmacoepidemiology study to compare the risk of MACE among migraine patients initiating aCGRP treatment or lisinopril/candesartan versus topiramate. By examining medications with potentially opposing effects on neurogenic inflammation, we aimed to elucidate the underlying mechanisms of migraine associated cardiovascular risk.

## Methods

### Study design and participants

This retrospective cohort study utilized data from the U.S. Department of Veterans Affairs (VA) Headache Centers of Excellence Administrative Data Cohort.^15^ The source population included incident migraine patients between Oct 1, 2007 and Sep 30, 2023. From this population, we emulated two separate target trials analyzing patients who were (i) prescribed lisinopril/candesartan or (ii) aCGRP treatment to those receiving topiramate for migraine prevention between June 1, 2018 and September 30, 2024. This study was approved by the VA Connecticut Healthcare System Research & Development Committee and granted a waiver of informed consent (IRBNet# 1784787). The study design followed the PRINCIPLED guideline^16^, with reporting follows the STROBE guideline for cohort studies. A detailed statistical analysis plan is provided in *eSAP*.

Key protocol components of the target trials were outlined in the *Supplement Table 1*. Migraine patients were identified using International Classification of Diseases 9^th^ and 10^th^ versions codes. Exclusion criteria included prior use of study medications before migraine diagnosis, use within the past 3 months, or no primary care visit in the past 12 months. For the target trial comparing lisinopril/candesartan with topiramate, patients with prior use of other ACEIs (e.g., enalapril) or ARBs (e.g., losartan), or with concurrent prescriptions of topiramate and any ACEI or ARB, were excluded. Similarly, for the target trial comparing aCGRP versus topiramate, patients with concurrent abortive/preventive aCGRP and topiramate were excluded. We emulated two separate two-arm trials–one comparing aCGRP with topiramate and other comparing lisinopril/candesartan with topiramate–instead of a single three-arm trial to preserve sample size. This approach allowed us to retain patients who were prescribed both aCGRP treatment and an ACEI or ARB, since prescribing aCGRP agents in the VA typically requires failure of at least three preventive medications (including ACEIs/ARBs) per VA Formulary Advisor. All treatment episodes that met the eligibility criteria were included, with baseline defined as the date of the first prescription dispensed.

### Data sources

The primary data source was the VA Corporate Data Warehouse (CDW) which contains structured electronic health records, including patient demographics, vital status, inpatient and outpatient encounters, laboratory, pharmacy dispensing data and provider information since October 1, 1999. To capture healthcare occurred outside the VA, we included Medicare and Medicaid claims from October 1, 2007, to December 31, 2022, and VA Community Care Data up to September 30, 2024. Cause of death data were not included, as they were only available through December 31, 2021.

### Treatment strategies

Details of medications are provided in *Supplement Table 2*. Topiramate, a migraine preventive agent that has not demonstrated effects on cardiovascular events^17^ was used as an active comparator. Galcanezumab (100 mg/mL) for cluster headache, topiramate/phentermine for weight management, and lisinopril/candesartan combinations (e.g., lisinopril and hydrochlorothiazide indicated for hypertension) were not included. Rimegepant for migraine prevention was defined as ≥12 tablets dispensed per 30 days. We defined adherence as receiving a refill within 90 days after the end of supply. The medication possession ratio (MPR) was calculated by summing the days’ supply of all prescriptions, dividing by the number of days from baseline to the end of treatment and truncating at the maximum value of 1.0.

### Outcomes

The primary outcome was MACE, defined as a composite of MI, ischemic stroke, intracerebral and subarachnoid hemorrhage (ICH/SAH), and all-cause mortality. Secondary outcomes were MI, ischemic stroke, ICH/SAH, respectively. Patients were followed from the baseline until the occurrence of an event, 60 months after the baseline, or administrative end of the study by September 30, 2024, whichever happened first.

### Covariates

We used a priori selection strategy^18^ to identify potential confounders based on subject-matter knowledge, literature and a directed acyclic graph (*Supplement Figure 1a*). We assumed treatment strategies were exchangeable conditional on baseline covariates, including demographics, social determinants of health, healthcare-seeking behavior, cardiovascular risk factors and related medications, migraine severity and treatments (*Supplement Table 3*). Missing data for continuous covariates were imputed with median values, and categorical variables with missing data were coded as “Unknown”.

### Statistical analysis

Baseline characteristics were summarized as frequencies and percentages, means and standardized deviations (SDs), or medians and interquartile ranges (IQRs) for non-normally distributed variables. Both intention-to-treat and per-protocol analyses were performed. Our approach to interpreting data is based on an evaluation of the magnitude, direction, and precision of the effect estimates rather than binary significance testing.^19^ Data were managed using SQL Server Management Studio 19.0, and statistical analyses were conducted using R version 4.3.1. All SQL queries and analytical code are available at https://github.com/kw375/aCGRP_CV/.

The intention-to-treat analyses examined the effect of initiating lisinopril/candesartan or aCGRP treatments versus topiramate on the risk of MACE. Observational analogs of the intention-to-treat effect were estimated by adjusting for baseline confounders using stabilized inverse probability of treatment initiating weights (IPTiW) via logistic regression models (details in eSAP). Covariate balance was assessed using standardized mean difference (SMD), with a value less than 0.1 indicating good balance between comparisons. Next, cumulative incidence curves were estimated using IPTiW-weighted pooled logistic regressions, with an indicator for treatment strategy, a flexible time-varying intercept, and a treatment-by-time interaction term. Five-year risk differences (RDs) and risk ratios (RRs) were compared. Confidence intervals (CIs) for cumulative incidence rates, RDs, and RRs were computed nonparametrically using 500 bootstrapping samples. The overall hazard ratios (HRs) over five years were obtained using pooled logistic regressions without the interaction term, with robust sandwich estimator for standardized errors.

For per-protocol analyses, observations not adherent to their initial treatment strategies were artificially censored. Stabilized inverse probability of treatment adherence weights (IPTaW) were estimated to account for potential selection bias associated with non-adherence, using logistical regression models additional adjusted for time-updated covariates, including body mass index, blood pressures, hospitalizations, diagnoses of chronic kidney disease (CKD), heart failure (HF), hypertension or chronic migraine, and medications (β-blockers, diuretics, antiplatelets, triptans, and non-steroid anti-inflammatory drugs [NSAIDs]). Final weights were calculated as the product of IPTiW and IPTaW. Due to computational constraints, for per-protocol analyses, only overall HRs were estimated via weighted pooled logistic regression.

To assess the robustness of intention-to-treat estimates, we conducted the following sensitivity analyses. (1) To address potential confounding by indication^20^, patients without documented cardiovascular indications (CKD, HF, hypertension, or history of MI) were excluded in the lisinopril/candesartan target trial, and (2) further stratified by baseline age and systolic blood pressure (SBP). (3) We repeated the primary analyses using outcome regressions to estimate the overall HRs, adjusting for the exact set of baseline confounders used in the IPTiW models, (4) and additionally estimated the sub-distribution hazards to account for competing risks from death for secondary outcomes.

We also conducted three bias assessments to detect the presence and/or quantify the magnitude of uncontrolled confounding. We (1) computed E-values^21^ based on the estimation from outcome regressions to quantify the magnitude unmeasured confounding have to be to negate the observed associations, and then (2) conducted probabilistic bias analyses specified in the *Supplement Methods* to assess the impact of residual confounding. (3) To detect spurious associations arising from selection bias, misclassification, residual confounding, and statistical analysis, we repeated the analyses on both positive and negative outcome controls. Given the well-documented efficacy of topiramate for weight loss^22^ and findings from the Food and Drug

Administration Adverse Event Reporting System^23^, we used weight change and primary malignancy as positive control outcomes. Influenza and non-pathological fracture, which have no known etiologic or pathophysiologic links to lisinopril/candesartan and aCGRP treatment, were selected as negative control outcomes^24, 25^ to assess residual confounding from healthcare-seeking behavior and frailty. Influenza diagnosis depends on vaccination uptake and symptom-driven testing, making it a proxy for healthcare-seeking behavior. Similarly, non-pathological fractures are often associated with frailty, which in turn is associated with cardiovascular events.

## Results

The study included 48,610 patients who initiated lisinopril/candesartan versus 115,047 patients who initiated topiramate, and 25,635 patients who initiated aCGRP treatment versus 135,740 patients who initiated topiramate (*Table 1 & Figure 1*).

**Table 1.**
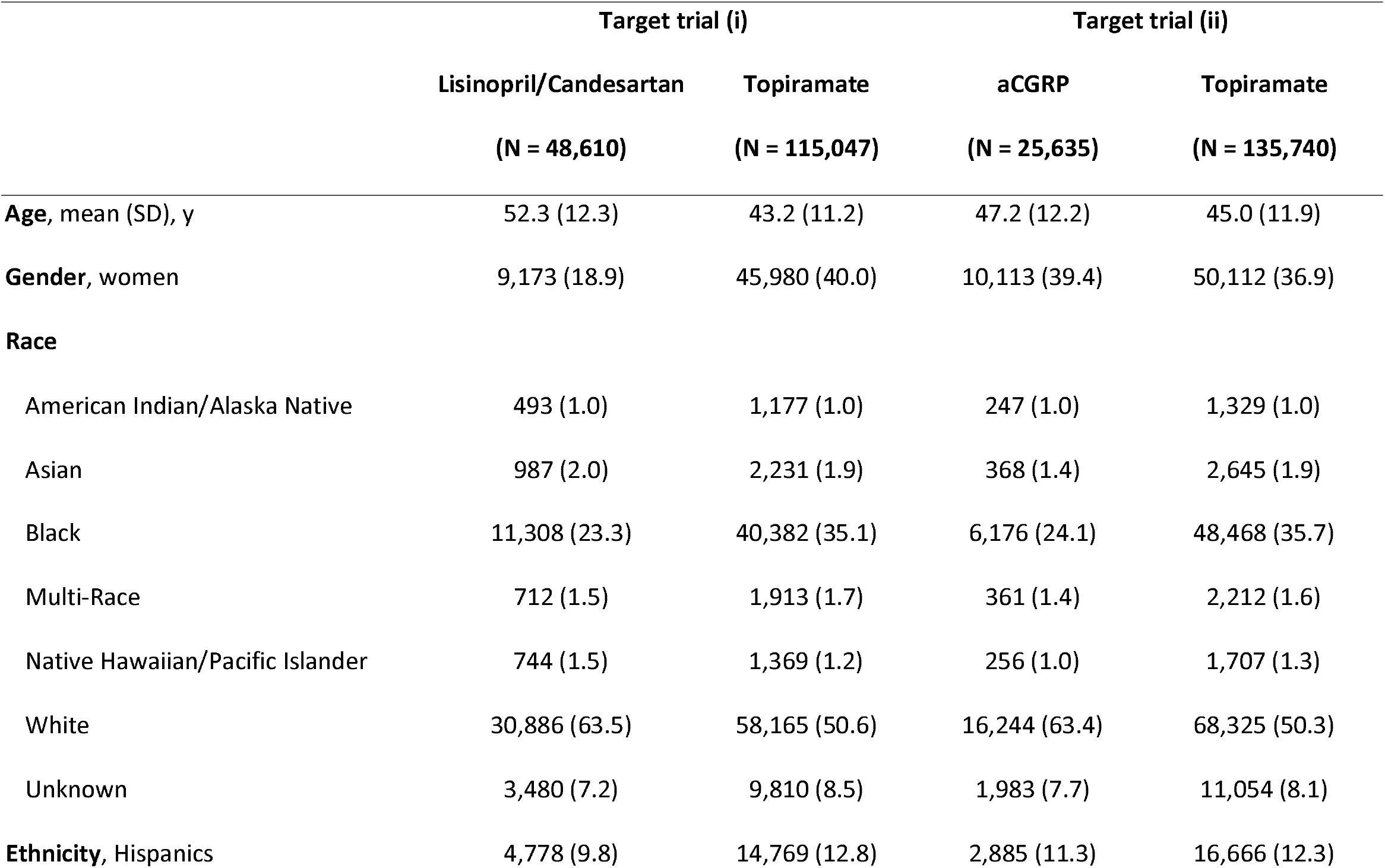
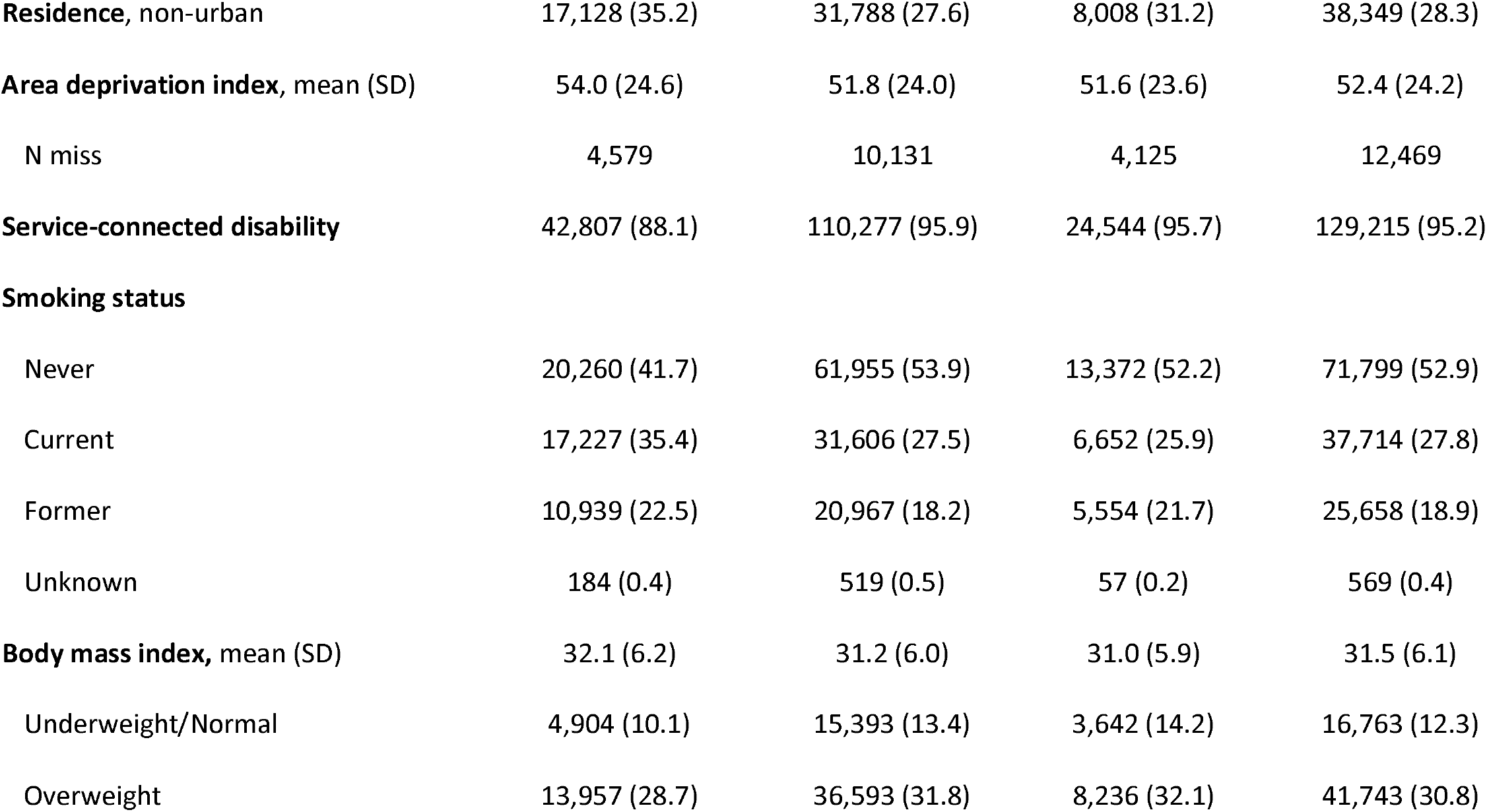
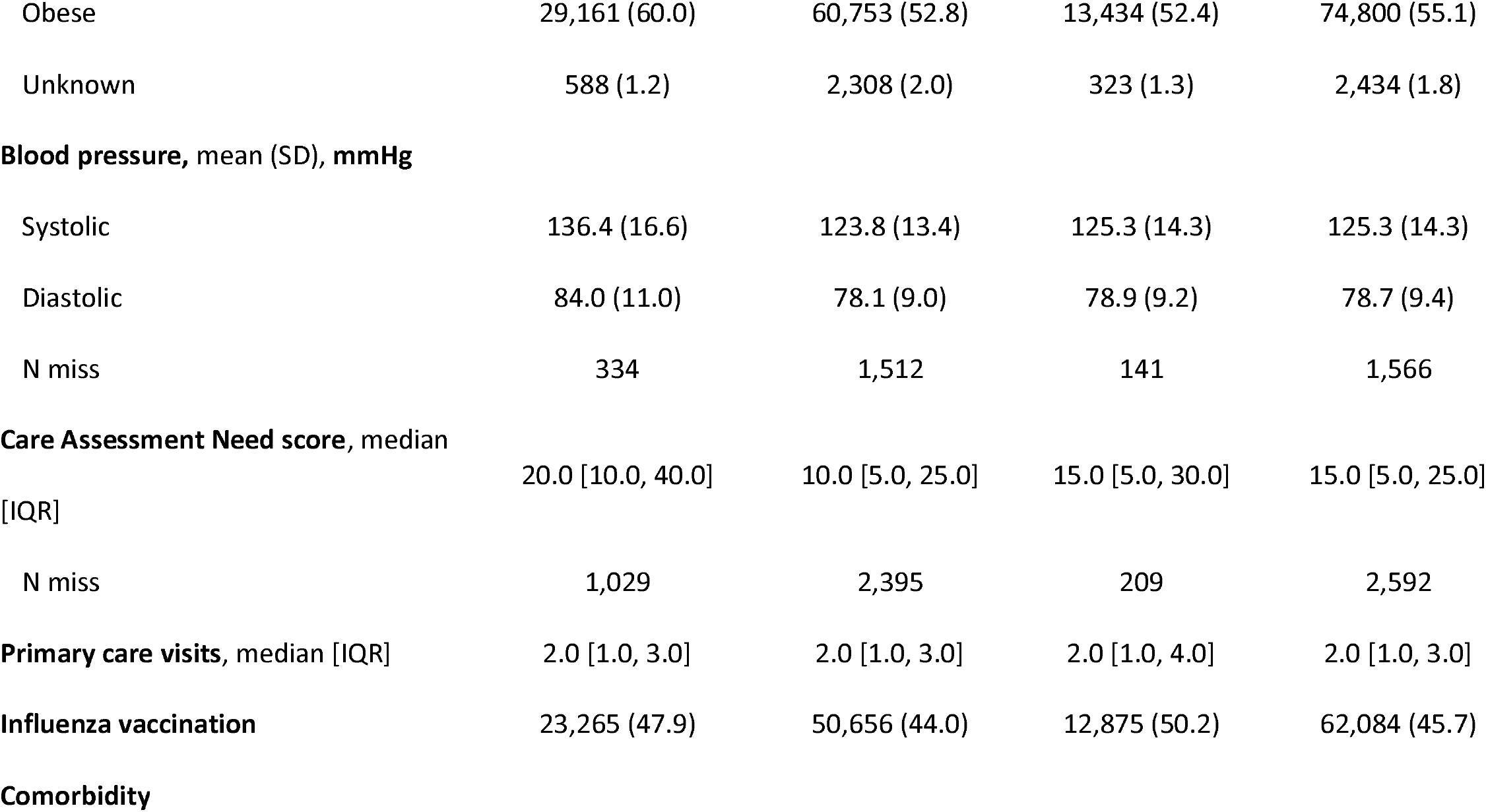
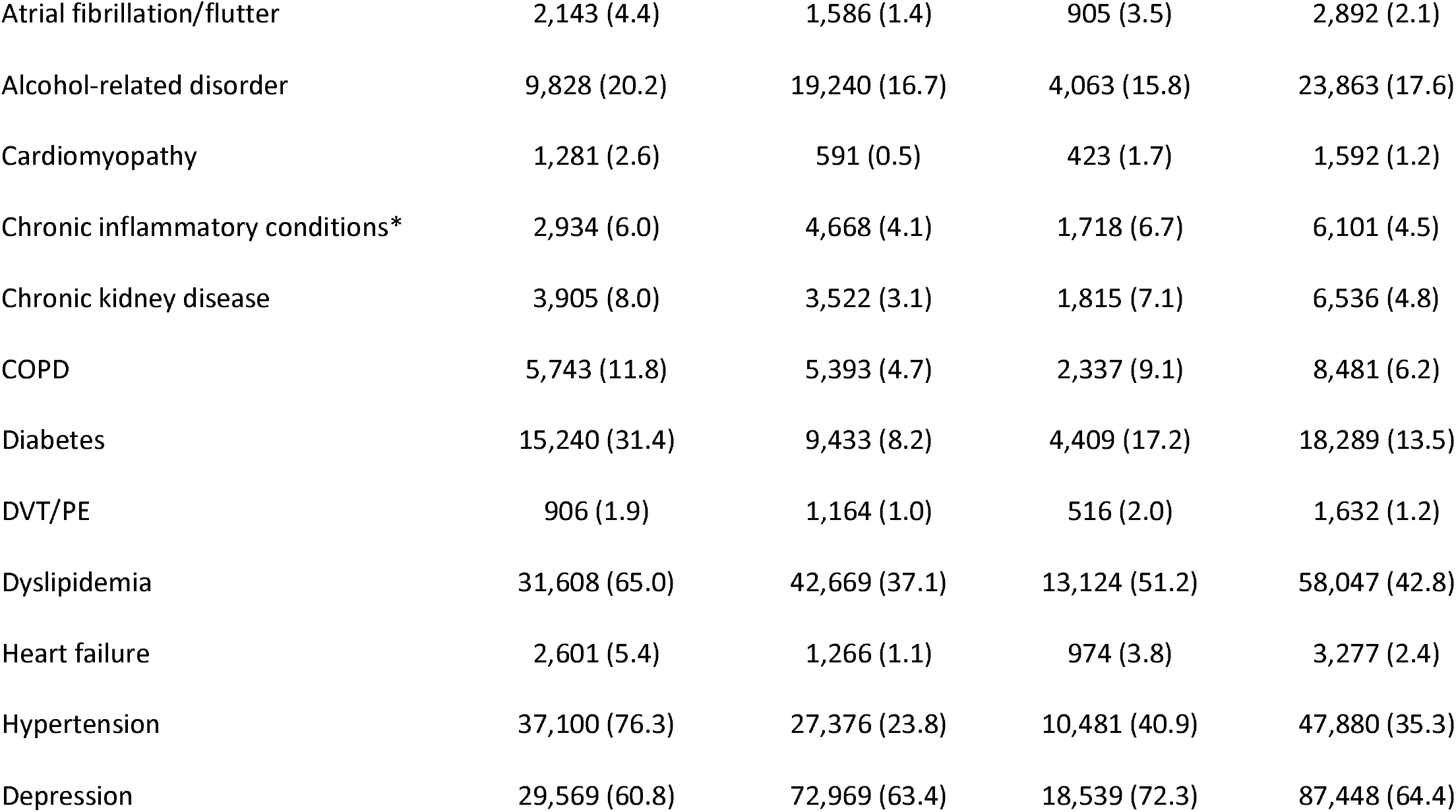
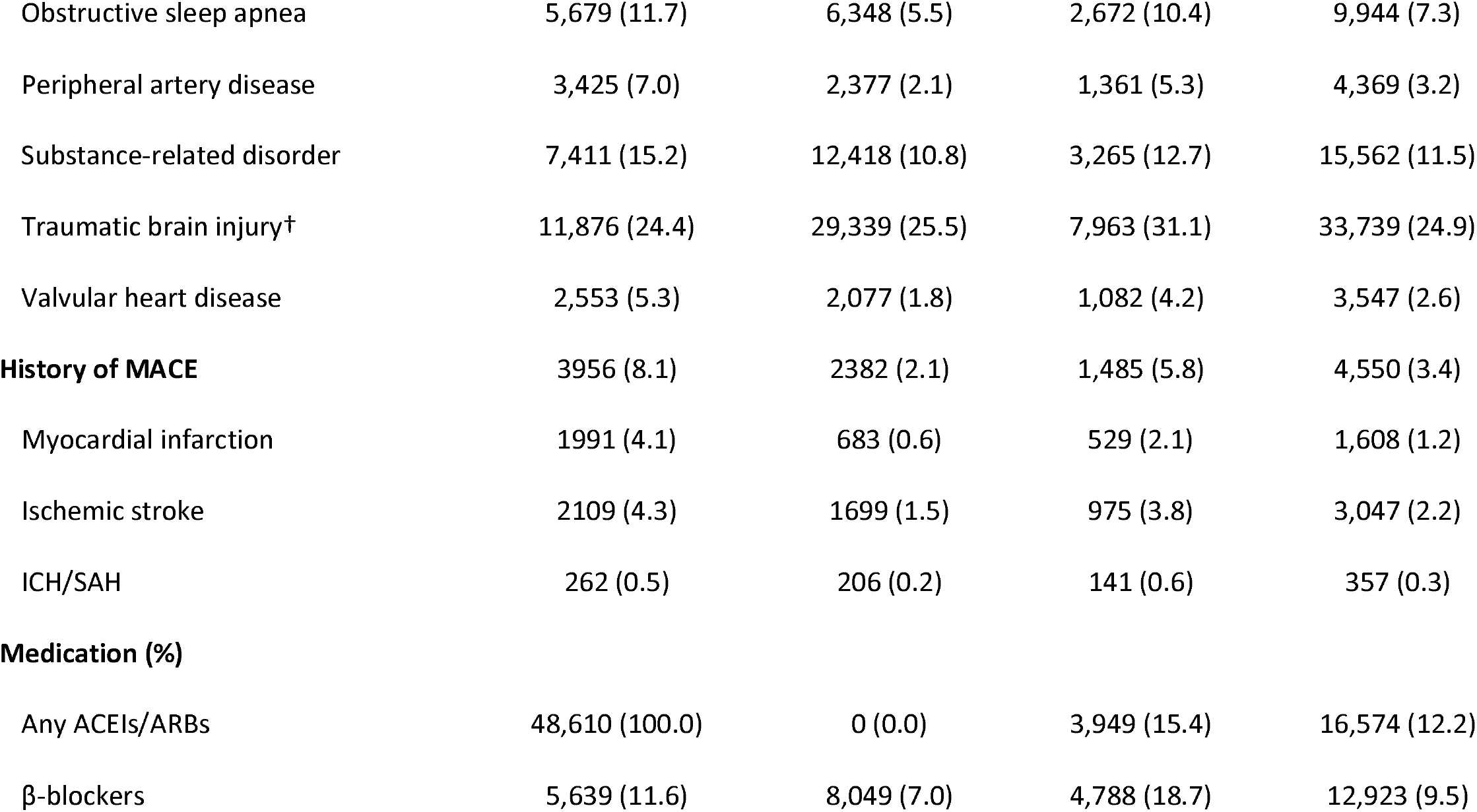
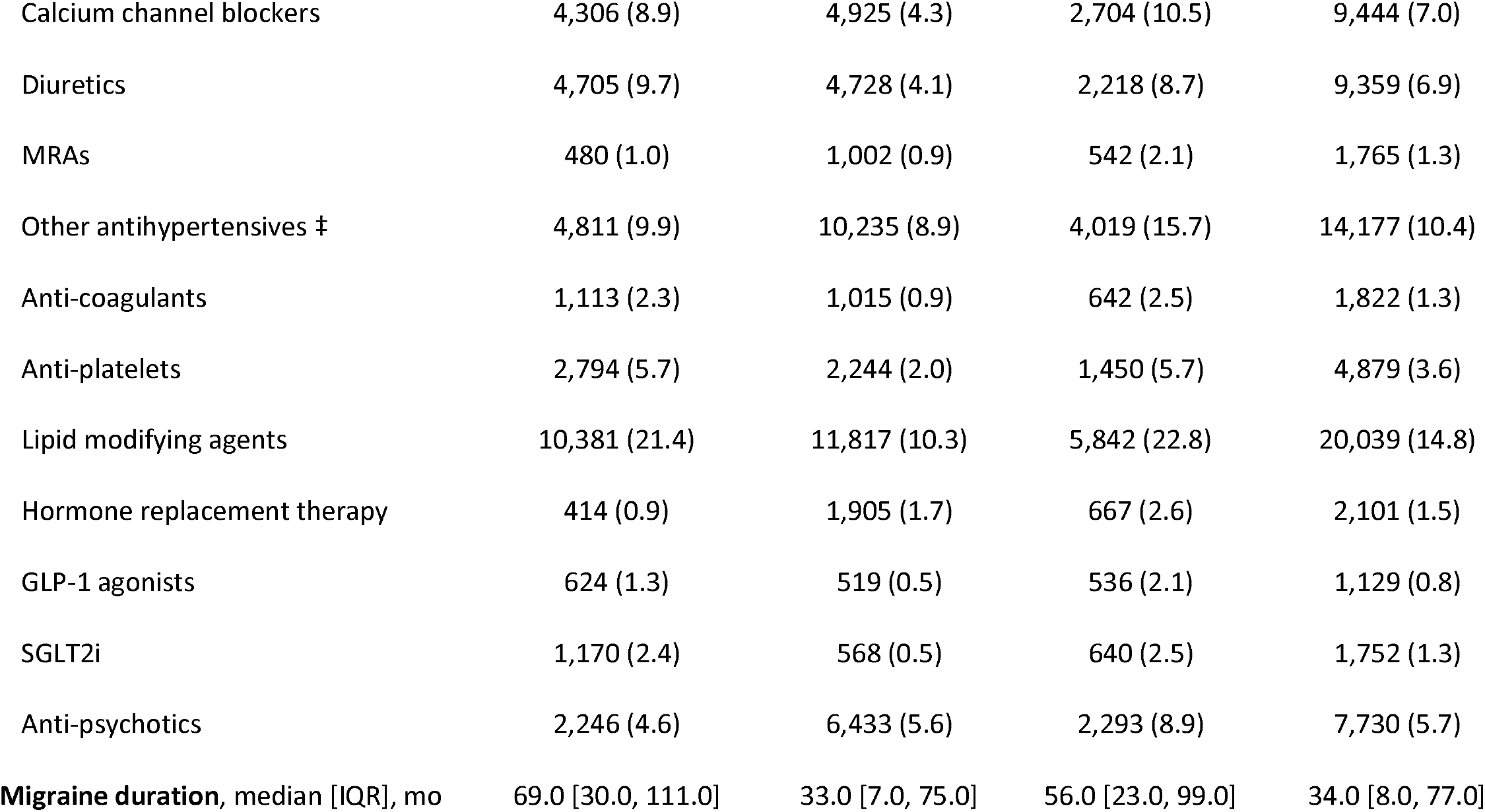
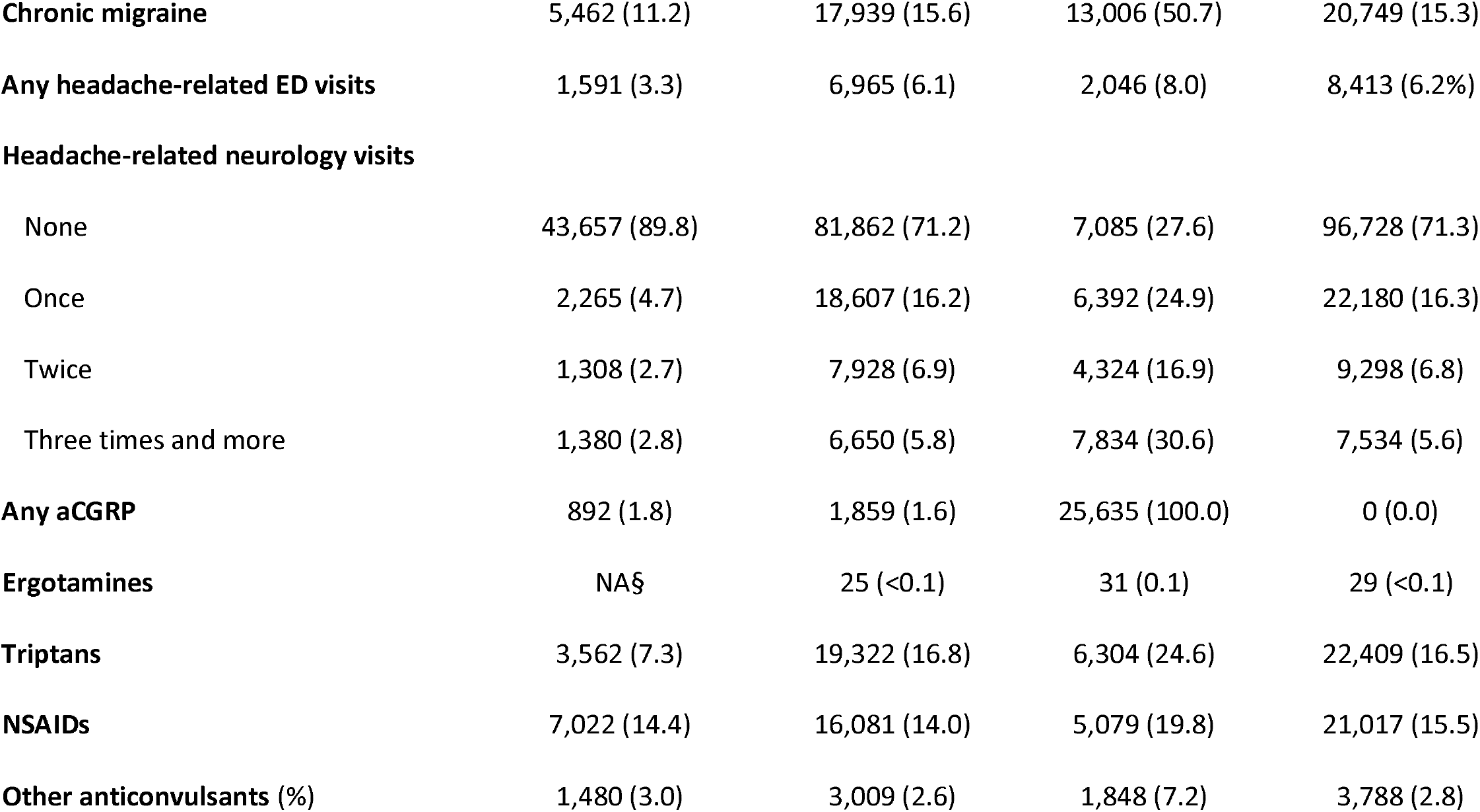
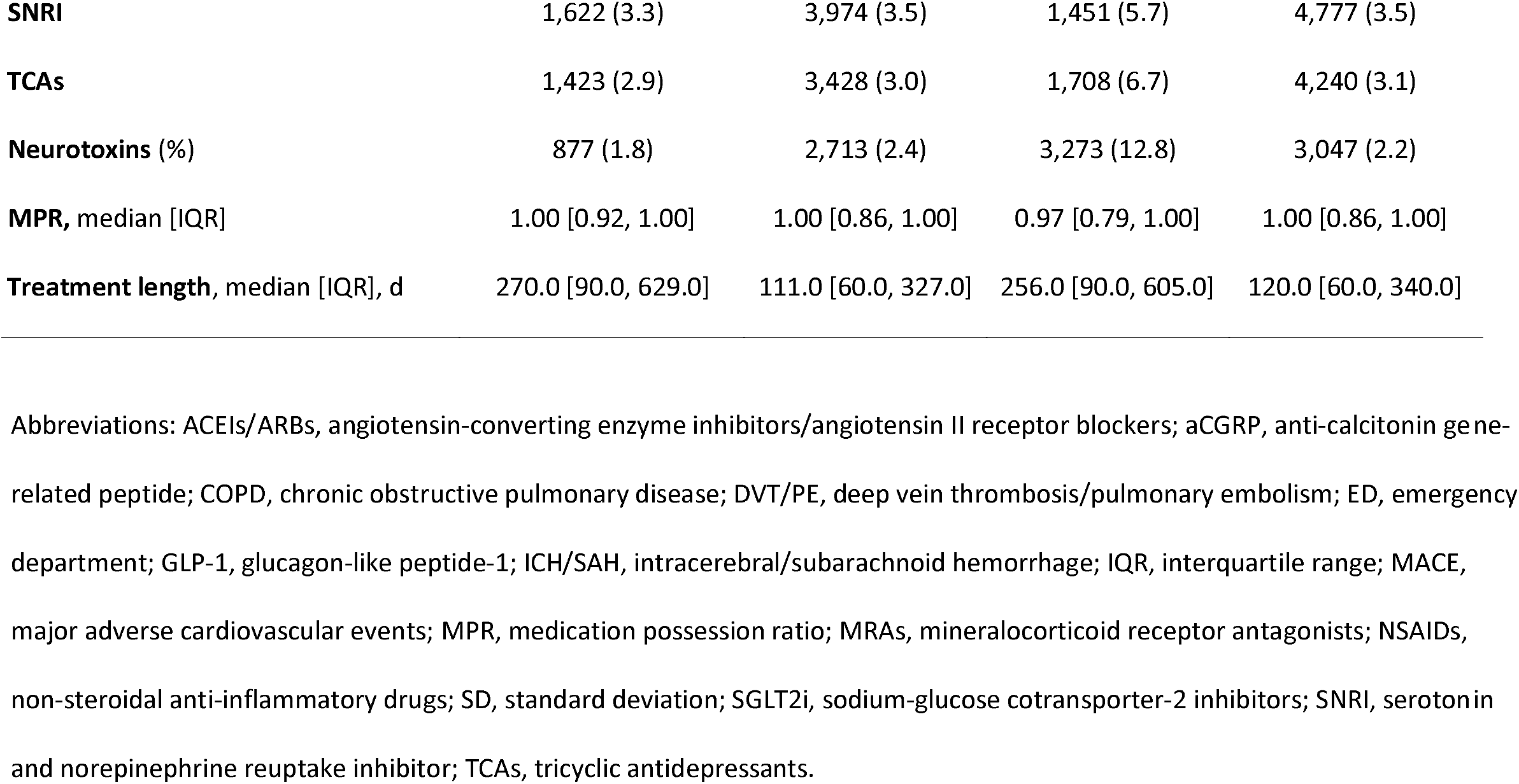
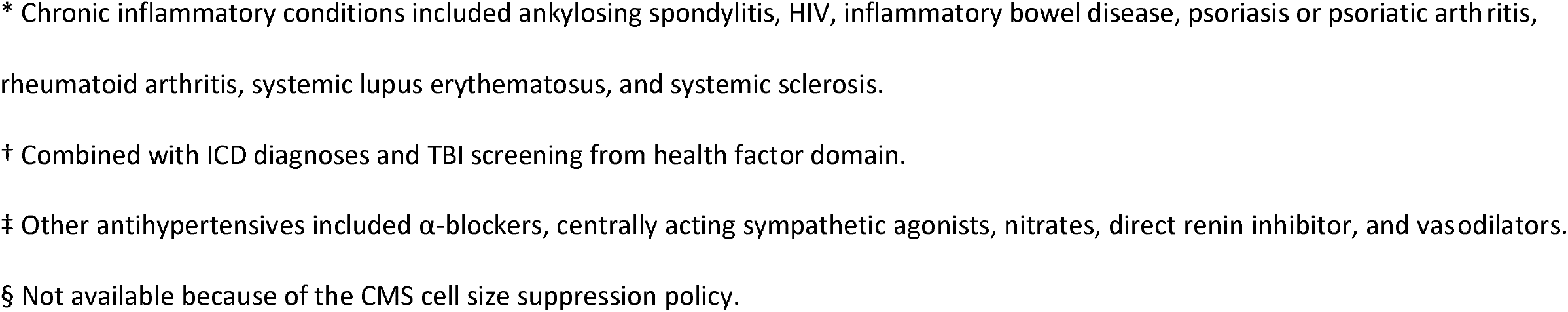
Baseline Characteristics of Person-Trials in the Study Sample.

**Figure 1.**
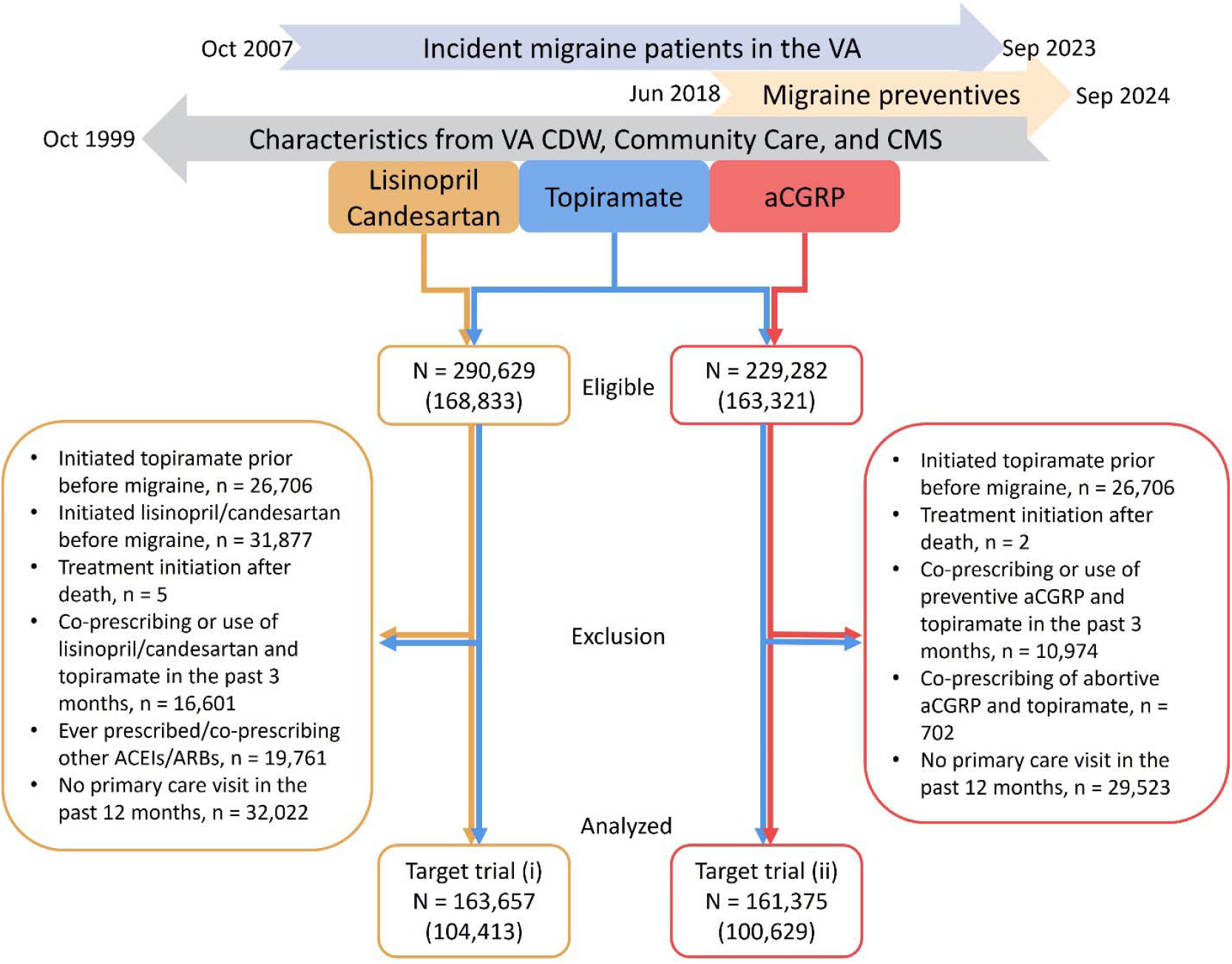
Study Flowchart. Numbers in the parentheses indicate the number of unique patients. ACEIs/ARBs indicates angiotensin-converting enzyme inhibitors/angiotensin II receptor blockers; aCGRP, anti-calcitonin gene-related peptide; CDW, Corporate Data Warehouse; CMS, Centers for Medicare & Medicaid Services; VA, Department of Veterans Affairs.

### Lisinopril/Candesartan versus Topiramate Target Trial

Patients initiating lisinopril/candesartan were older (mean [SD], 52.3 [12.3] vs. 43.2 [11.2] years), with fewer women (18.9% vs. 40.0%), Black (23.3% vs. 35.1%) and Hispanic individuals (9.8% vs. 12.8%). These patients had higher SBP (136.4 [16.6] vs. 123.8 [13.4] mm Hg) and cardiovascular risks at baseline, including hypertension (76.3% vs. 23.8%), diabetes (31.4% vs. 8.2%), dyslipidemia (65.0% vs. 37.1%), and history of MI (4.1% vs. 0.6%) or ischemic stroke (4.3% vs. 1.5%). Use of antiplatelets (5.7% vs. 2.0%), β-blockers (11.6% vs. 7.0%), and lipid-modifying agents (21.4% vs. 10.3%) was also higher, while chronic migraine diagnosis (11.2% vs. 15.6%) and triptan use (7.3% vs. 16.8%) were less frequent. Notably, comorbid conditions commonly managed by primary care, such as hypertension and dyslipidemia, were well captured in the VA CDW data, while cardiovascular events were more accurately captured using VA Community Care Data (*Supplement Table 4*).

In the intention-to-treat analysis, the median [IQR] follow-up time was 35.0 [20.0-55.0] months for lisinopril/candesartan and 39.0 [23.0-57.0] months in the topiramate. After applying IPTiW, baseline covariates were well balanced (*Supplement Figure 2*). The 5-year cumulative incidence (*Figure 2*) of MACE was 7.08% (95% CI, 6.57-7.64%) in the lisinopril/candesartan group versus 6.01% (5.54-6.54%) in the topiramate group, which results in a RD of 1.06% (0.35-1.85%), a RR of 1.18 (1.05-1.33), and an average HR of 1.21 (1.10-1.34) by 5 years (Table 2).

**Table 2.**
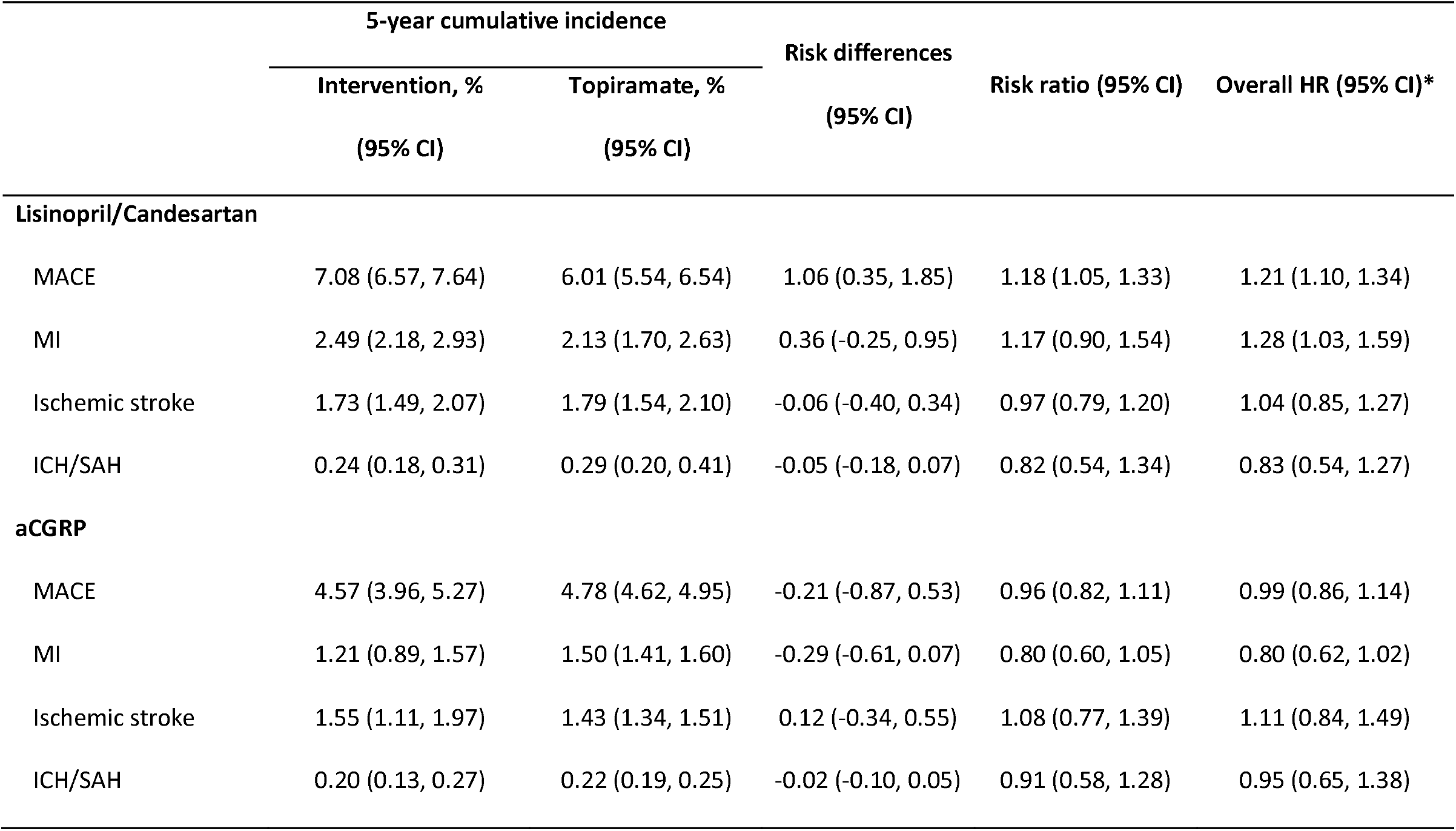
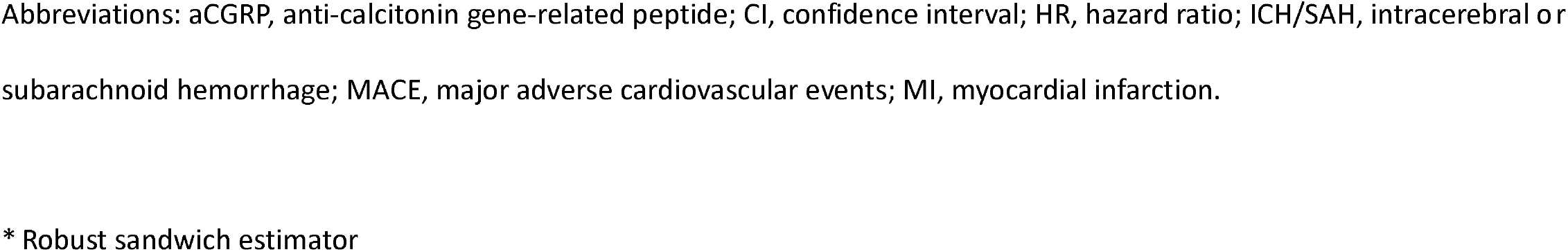
Intention-to-treat estimates of cumulative incidence, risk differences (RD), risk ratio (RR) and overall hazard ratio (HR) for major adverse cardiovascular events and secondary outcomes over 5 years.

|  | 5-year cumulative incidence |  | Risk differences<br>(95% CI) | Risk ratio (95% CI) | Overall HR (95% CI)* |
| --- | --- | --- | --- | --- | --- |
|  | Intervention, % | Topiramate, % |  |  |  |
|  | (95% CI) | (95% CI) |  |  |  |
| Lisinopril/Candesartan |  |  |  |  |  |
| MACE | 7.08 (6.57, 7.64) | 6.01 (5.54, 6.54) | 1.06 (0.35, 1.85) | 1.18 (1.05, 1.33) | 1.21 (1.10, 1.34) |
| MI | 2.49 (2.18, 2.93) | 2.13 (1.70, 2.63) | 0.36 (-0.25, 0.95) | 1.17 (0.90, 1.54) | 1.28 (1.03, 1.59) |
| Ischemic stroke | 1.73 (1.49, 2.07) | 1.79 (1.54, 2.10) | -0.06 (-0.40, 0.34) | 0.97 (0.79, 1.20) | 1.04 (0.85, 1.27) |
| ICH/SAH | 0.24 (0.18, 0.31) | 0.29 (0.20, 0.41) | -0.05 (-0.18, 0.07) | 0.82 (0.54, 1.34) | 0.83 (0.54, 1.27) |
| aCGRP |  |  |  |  |  |
| MACE | 4.57 (3.96, 5.27) | 4.78 (4.62, 4.95) | -0.21 (-0.87, 0.53) | 0.96 (0.82, 1.11) | 0.99 (0.86, 1.14) |
| MI | 1.21 (0.89, 1.57) | 1.50 (1.41, 1.60) | -0.29 (-0.61, 0.07) | 0.80 (0.60, 1.05) | 0.80 (0.62, 1.02) |
| Ischemic stroke | 1.55 (1.11, 1.97) | 1.43 (1.34, 1.51) | 0.12 (-0.34, 0.55) | 1.08 (0.77, 1.39) | 1.11 (0.84, 1.49) |
| ICH/SAH | 0.20 (0.13, 0.27) | 0.22 (0.19, 0.25) | -0.02 (-0.10, 0.05) | 0.91 (0.58, 1.28) | 0.95 (0.65, 1.38) |

Abbreviations: aCGRP, anti-calcitonin gene-related peptide; CI, confidence interval; HR, hazard ratio; ICH/SAH, intracerebral or subarachnoid hemorrhage; MACE, major adverse cardiovascular events; MI, myocardial infarction.
\* Robust sandwich estimator

**Figure 2.**
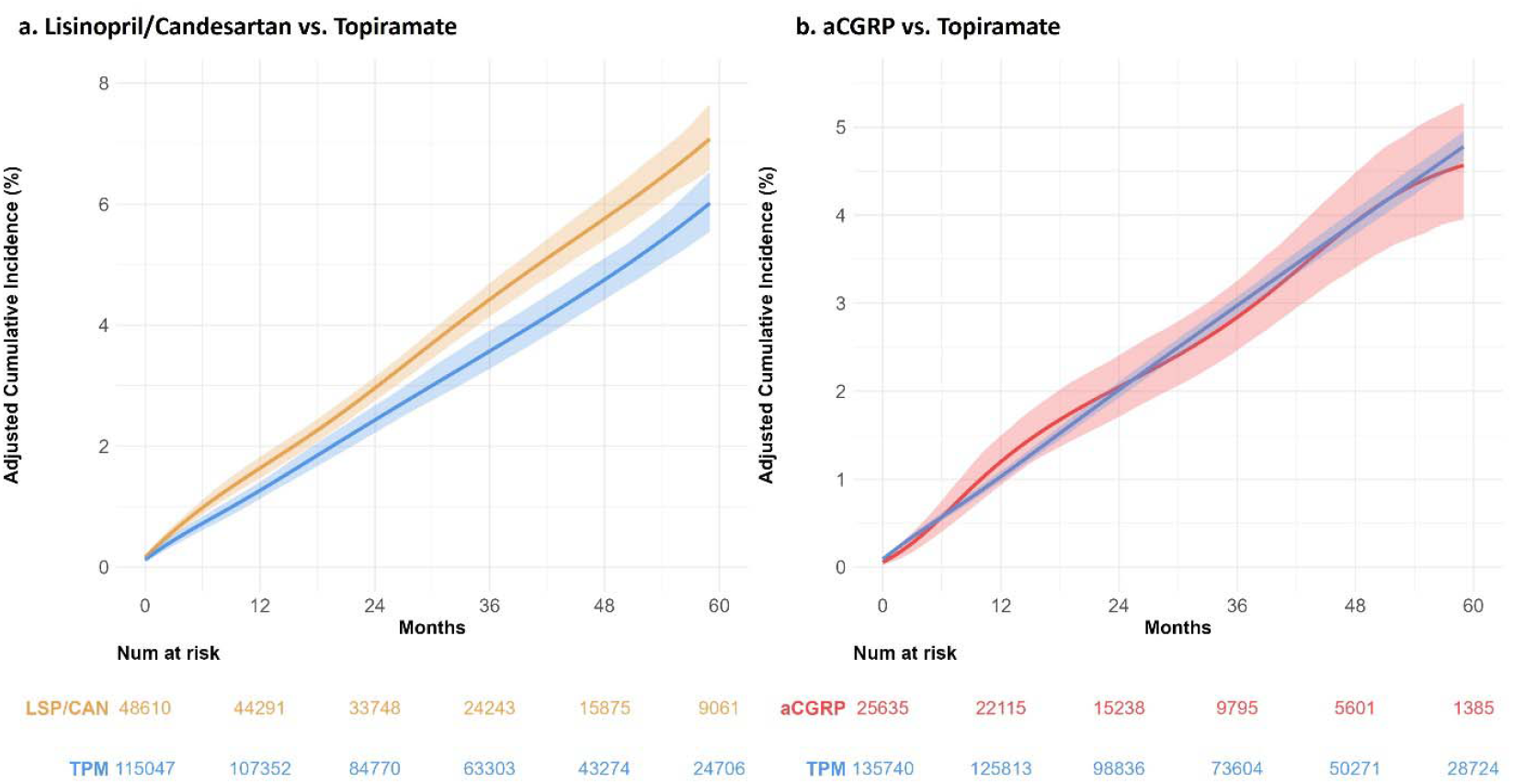
Intention-to-Treat Cumulative Incidence of Major Adverse Cardiovascular Events. Cumulative incidence from the target trial comparing (a) lisinopril/candesartan (LSP/CAN) vs topiramate (TPM); (b) Cumulative incidence from the target trial comparing anti-calcitonin gene-related peptide (aCGRP) treatment vs TPM. The shaded ribbons indicate 95% CIs based on 500 bootstrapping samples.

For secondary outcomes, the 5-year cumulative incidence after initiating lisinopril/candesartan was 2.49% (2.18-2.93%) for MI, 1.73% (1.49-2.07%) for ischemic stroke and 0.24% (0.18-0.31%) for ICH/SAH (*Figure 3*). A higher risk of MI was observed in the lisinopril/candesartan arm (RD 0.36% [-0.25-0.95%]; RR 1.17 [0.90-1.54]; and HR 1.28 [1.03-1.59]). The risk of ischemic stroke was similar in both groups (HR 1.04; 95% CI, 0.85-1.27), while the risk of ICH/SAH was lower (HR 0.83; 95% CI, 0.54-1.27) in the lisinopril/candesartan group.

**Figure 3.**
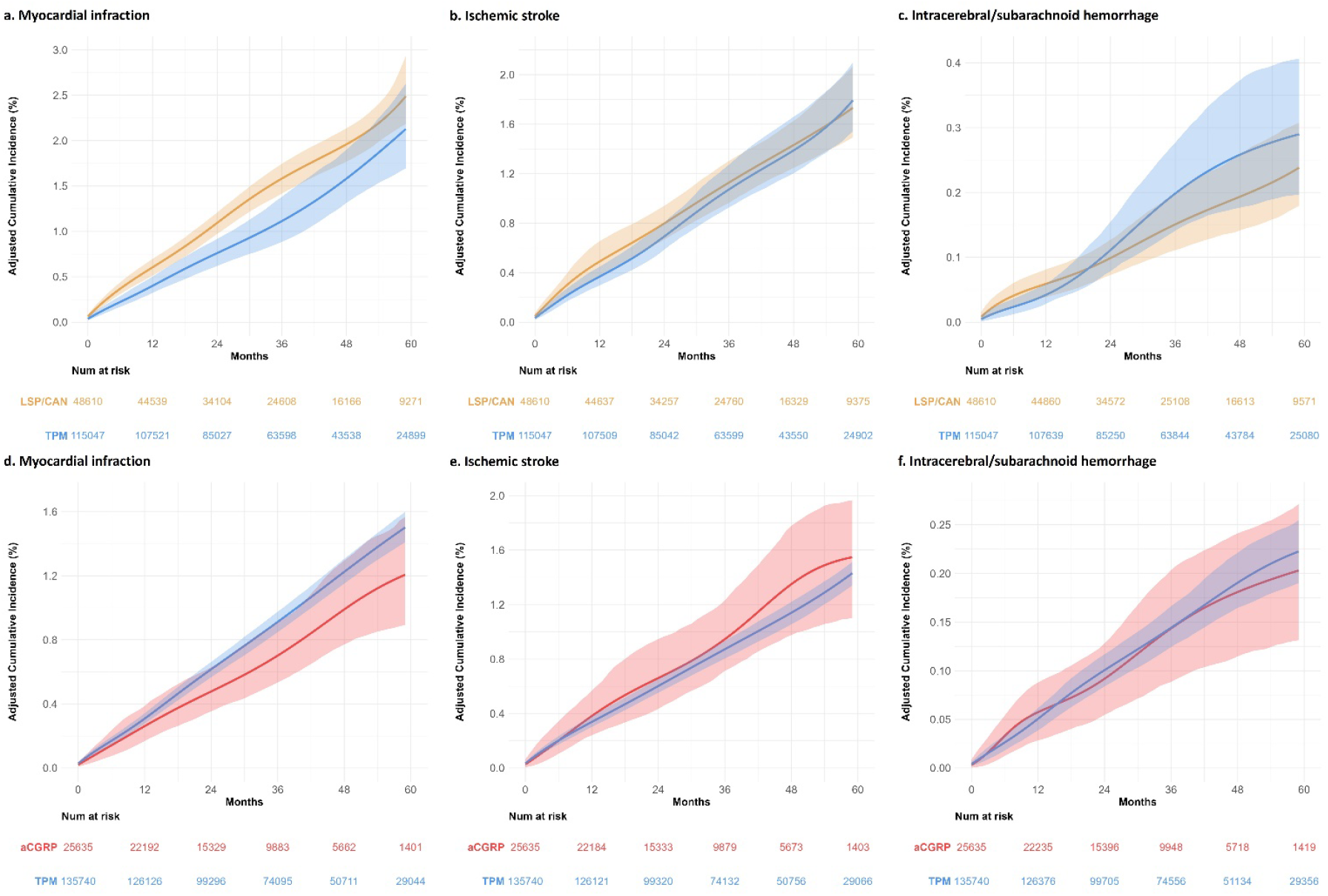
Intention-to-Treat Cumulative Incidence of Secondary Outcomes. Cumulative incidence of (a) myocardial infarction (MI), (b) ischemic stroke, and (c) intracerebral/subarachnoid hemorrhage (ICH/SAH) from the target trial comparing lisinopril/candesartan (LSP/CAN) versus topiramate (TPM); and (d) MI, (e) ischemic stroke, and (f) ICH/SAH from the target trial comparing anti-calcitonin gene-related peptide (aCGRP) treatment versus TPM. The shaded ribbons represent 95% CIs based on 500 bootstrapping samples.

In the per-protocol analysis, the median follow-up time was 8.0 [3.0-19.0] months for lisinopril/candesartan and 4.0 [2.0-11.0] months for the topiramate. The mean IPTaW was 1.000 and ranged from 0.028 to 26.747 (*Supplement Table 5*). The risk of MACE (HR 1.30; 95% CI, 1.09-1.58) and MI (HR 1.52; 95% CI, 1.07-2.17) remained higher in the lisinopril/candesartan group (Table 3).

**Table 3.** Per-protocol estimates of overall hazard ratio (HR) of major adverse cardiovascular events and secondary outcomes over 5 years, after censoring individuals who did not adhere to the treatment.

|  | **Lisinopril/Candesartan** | **aCGRP** |
| --- | --- | --- |
|  | HR (95% CI)* | HR (95% CI)* |
| **MACE** | 1.30 (1.09, 1.58) | 0.92 (0.70, 1.21) |
| MI | 1.52 (1.07, 2.17) | 0.71 (0.45, 1.14) |
| Ischemic stroke | 1.16 (0.87, 1.55) | 1.00 (0.58, 1.73) |
| ICH/SAH | 1.71 (0.90, 3.23) | 0.73 (0.37, 1.46) |

Abbreviations: aCGRP, anti-calcitonin gene-related peptide; CI, confidence interval; HR, hazard ratio; ICH/SAH, intracerebral/subarachnoid hemorrhage; MACE, major adverse cardiovascular events; MI, myocardial infarction.
\* Robust sandwich estimator

### aCGRP versus Topiramate Target Trial

The aCGRP and topiramate groups had similar age (47.2 [12.2] vs. 45.0 [11.9] years) and proportion of women (39.4% vs. 36.9%), but a lower proportion of black individuals (24.1% vs. 35.7%). Both groups had comparable SBP (125.3 [14.3] mm Hg), although the aCGRP group had slightly higher cardiovascular risks, including hypertension (40.9% vs. 35.3%), history of MI (2.1% vs. 1.2%) and ischemic stroke (3.8% vs. 2.2%). The aCGRP group also had more severe migraine characteristics, such as traumatic brain injury (31.1% vs. 24.9%), chronic migraine (50.7% vs. 15.3), at least one headache-related neurology visits (72.4% vs. 28.7%), and higher triptan (24.6% vs. 16.5%) and neurotoxin use (12.8% vs. 2.2%).

In the intention-to-treat analysis, median [IQR] follow-up was 28.0 [16.0-45.0] months for aCGRP and 38.0 [22.0-57.0] for topiramate. The 5-year cumulative incidence of MACE was 4.57% (3.96-5.27%) in the aCGRP group and 4.78% (3.96-5.27%) in the topiramate group. The risks of MACE (HR 0.99; 95% CI, 0.86-1.14), ischemic stroke (HR 1.11; 95% CI, 0.94, 1.49) and ICH/SAH (HR 0.95; 95% CI, 0.65-1.38) were similar between groups. However, the risk of MI appeared lower in the aCGRP group (cumulative incidence: 1.21% vs. 1.50%), with a 5-year RD of −0.29% (−0.61-0.07%), RR of 0.80 (0.60-1.05), and an average HR of 0.80 (0.62-1.02).

In the per-protocol analysis, median follow-up time was 7.0 [3.0-17.0] months for aCGRP and 4.0 [2.0-11.0] for topiramate. The mean IPTaW was 1.001 and ranged from 0.021 to 42.704. The risk of MACE remained similar (HR 0.92; 95% CI, 0.70-1.21) between groups, and the risk of MI was also lower in the aCGRP arm (HR 0.71; 95% CI, 0.45-1.14).

### Sensitivity Analyses and Bias Assessments

Results of the sensitivity analyses are presented in *Supplement Tables 6-10*. Excluding patients with documented cardiovascular indications for lisinopril/candesartan yielded HRs of 1.70 (95% CI, 1.44-2.03) for MACE and 2.75 (95% CI, 2.01-3.77) for MI. Stratification of the overall sample (*Supplement Figure 3*) showed higher risks of MACE (HR 1.63; 95% CI, 1.16-2.29) and MI (HR 3.18; 95% CI, 1.63-6.20) in younger patients < 40 years old, or those with SBP < 130 (MACE 1.63, 95% CI, 1.43-1.86; MI 1.86, 95% CI, 1.44-2.39) and between 130-139 mm Hg (MACE 1.16, 95% CI, 1.00-1.34; MI 1.46, 95% CI, 1.11-1.92). The increased risks of MACE and MI among younger patients or those with SBP < 140 mm Hg were more pronounced among those without cardiovascular indications for lisinopril/candesartan. In contrast, the risks of MACE (HR 0.79; 95% CI, 0.65-0.97) and ischemic stroke (HR 0.64; 95% CI, 0.47-0.88) were lower in the overall sample of patients with SBP ≥ 140 mm Hg. After accounting for competing risks from death, the lisinopril/candesartan group showed higher risks of MI (HR 1.48; 95% CI, 1.31-1.67). Conversely, the aCGRP group showed a lower risk of MI (HR 0.82; 95% CI, 0.70-0.97).

The E-values (*Supplement Table 8*) were 1.81 for the association of lisinopril/candesartan on MACE, and 2.32 on MI, when compared to topiramate. For the comparison of aCGRP versus topiramate, the E-value for MI was 1.74. In the probabilistic bias analyses (*Supplement Table 9*), the association between aCGRP treatment and MI was attenuated after adjustment for potential unmeasured confounder: monthly migraine days (RR 0.88; 95% simulation interval [SI], 0.62 – 1.19), monthly headache days (RR 0.92; 95% SI, 0.71 – 1.18), or migraine with aura (RR 0.90; 95% SI, 0.69 – 1.16). For the association between lisinopril/candesartan on the risk of MI among individuals without documented cardiovascular indications, bias-adjusted RRs excluded null under scenarios with moderate to strong unmeasured confounding, and only when assuming an unknown cardiovascular risk as strong as a history of MI, did the 95% SI cross the null (RR, 1.45; 95% SI, 0.90 – 2.14). after adjustment for strong unknown confounding using a hypothetical cardiovascular risk factor, the bias-adjusted RRs excluded the null, only when an unknown cardiovascular risk factor as strong as history of MI, the 95% SI crossed 1 (RR 1.45; 95% SI 0.90 – 2.14). For outcome controls, both lisinopril/candesartan and aCGRP groups had higher risks of primary malignancy and weight increase compared to topiramate. The risk of non-pathological fracture was comparable in the lisinopril/candesartan group (HR 1.06; 95% CI, 0.98-1.14) but elevated in the aCGRP group (HR 1.13; 95% CI, 1.01-1.26). The risks of influenza were similar between comparisons.

## Discussion

In this retrospective study, we emulated two separate target trials to evaluate the association of lisinopril/candesartan and aCGRP treatments, compared to topiramate, for migraine prevention with the risk of MACE. We found that lisinopril/candesartan use was associated with a higher risk of MACE, primarily driven by MI, in the overall sample. This risk was greater in younger patients or those without documented cardiovascular indications or SBP <140 mmHg. However, the risk of ischemic stroke among individuals with SBP ≥ 140 mm Hg was 36% lower, which is consistent with the guideline recommendation to initiate ACEIs/ARBs to prevent stroke in hypertensive adults.^26^ Meanwhile, aCGRP treatment was associated with a reduced risk of MI, and was not associated with ischemic stroke or ICH/SAH. Lastly, we did not observe associations of lisinopril/candesartan and aCGRP treatment on the risk of ICH/SAH, possibly due to low incident rate of ICH/SAH and insufficient power.

The observation that lisinopril/candesartan may increase cardiovascular risk appears to contradict current clinical guidelines. However the evidence for cardiovascular benefits of ACEI/ARB is not specific to migraine patients. Migraine-associated cardiovascular risk may arise through mechanisms distinct from traditional atherosclerotic pathways. Two population-level observational studies have reported greater risk of cardiovascular events in younger migraine patients. For instance, a Danish study^27^ reported HRs for MI of 1.62 (95% CI, 1.33-1.98) in those aged 40 years or younger, 1.56 (1.38-1.77) in those aged 40-59 years, and 1.41 (1.21-1.64) in those 60 years or older. Similarly, a Korean study^28^ found HRs for stroke was 1.74 (1.65-1.84) for individuals under 60 years old, and 1.22 (1.18-1.26) for those aged 60 and older. These findings suggest that the cardiovascular risks associated with migraine are not primarily driven by atherosclerosis, which typically increase with age. Thus, while lisinopril/candesartan may reduce risks mediated by hypertension and atherosclerosis, they may not confer the same protection, and may potentially increase risk, in the context of migraine-driven, non-atherosclerotic mechanisms.

Although the mechanisms linking migraine-associated cardiovascular risk remain unclear, the release of CGRP and SP following trigeminal nerve activation may contribute via vasodilation and neurogenic inflammation. SP also promotes platelet aggregation and clots formation, and higher plasma SP levels have been associated with recurrent infarction or death by 2 years among post-MI patients.^29^ Inhibiting ACE, which normally degrades SP, may exacerbate neurogenic inflammation and increase MI risk, especially if ACE expression may be down-regulated in migraine patients.^30^ This SP-mediated pathway may also explain the increased cardiovascular risk observed in individuals with chronic pain^31, 32^, especially given ACEI use following a fracture has been associated with a higher risk of complex regional pain syndrome in a dose-dependent manner.^33^ In our sample predominantly with lisinopril (98.3%), the increased MI risk may reflect SP accumulation and neurogenic inflammation among individuals with minimal atherosclerotic risk (i.e., younger age, SBP < 140 mm Hg, and without documented CKD, HF, hypertension or history of MI). This inflammation may also affect cerebral vascular system, as the risk of ischemic stroke was higher in the lisinopril/candesartan group with SBP < 130 mm Hg (eTable 7, HR 1.56; 95% CI, 1.15-2.21). However, due to small sample size of younger patients or those with low atherosclerotic risks and relatively low incident rate of ischemic stroke, this association should be further validated in a larger cohort.

Current evidence from *in vitro* and animal studies suggests that CGRP may mediate a cardioprotective effect following myocardial injuries through its vasodilation, inotropy and chronotropy effects.^34^ However, the causal relationship of CGRP as the exposure has yet to be directly studied. To date, only two observational studies have compared the cardiovascular safety of aCGRP versus neurotoxin. One study, based on an underpowered Medicare sample, reported a HR of 0.86 (95% CI 0.30 - 2.48)^35^, but relied on machine learning-estimated propensity scores, which may yield biased estimates in causal inference^36^. Another insurance claim-based study reported a relative risk of 0.87 (95% CI 0.19 – 1.55) for MI.^37^ Despite their limitations, the direction of these point estimates aligns with our findings from the intention-to-treat analysis (HR 0.80; 95% CI 0.62-1.02). Although none of the three studies demonstrated statistically significant associations individually, had a meta-analysis pooled these estimates, we would likely observe a statistically significant association suggesting lower cardiovascular risk with aCGRP treatment. The smaller effect sizes observed in prior studies are likely attributable to differences in comparator selection. Neurotoxins are typically prescribed for patients with migraine (≥15 migraine days per month) who have failed multiple preventive treatments. As such, migraine frequency and severity may be more comparable between patients initiated with aCGRP and neurotoxin than between patients initiated with aCGRP and topiramate. In our probabilistic bias analysis adjusting for potential confounding by monthly migraine days, the bias-adjusted estimates (RR 0.88; 95% SI, 0.62 - 1.19) closely mirrored the estimates from neurotoxin comparisons. These findings are consistent with the hypothesis that migraine-associated cardiovascular risk may be mediated by neuroinflammatory process involving CGRP and SP released during migraine attacks. By blocking CGRP or its receptor, aCGRP treatment could reduce the long-term neurogenic inflammation in patients with migraine and thus lower the risk of MI.

This study has several limitations. The primary concern is confounding by indication, a type of confounding bias^38^ arises when the indication for treatment (disease or disease severity) is unmeasured or misclassified. The lisinopril/candesartan group was older, had higher SBP and had greater cardiovascular comorbidities, consistent with clinical practice where these agents are more frequently prescribed to migraine patients with or at risk of atherosclerotic disease. While β-blockers could be a closer equipoise active comparator, particularly if the increased risk of MACE/MI is driven by SP-related neuroinflammation, the positivity assumption in our analysis holds, as both lisinopril/candesartan and topiramate are indicated for migraine prevention. Although the precise indication, whether for migraine prevention, hypertension, or both, could not be ascertained with EHR data, conditional exchangeability remains plausible given extensive adjustment for atherosclerotic risk factors and other confounders. Besides, baseline covariates balances (SMDs) were achieved after weighting, large E-values which exceeded the HRs of known cardiovascular risk factors included in the model (*Supplement* Table 8), and findings from probabilistic bias analysis adjusting for strong unknown cardiovascular risk factors (*Supplement Table 9*), suggesting that the increased risk of MACE with lisinopril/candesartan is unlikely biased by residual confounding or unmeasured cardiovascular indications. Additionally, negative outcome control analyses for influenza (confounding by healthcare-seeking behavior) and non-pathological fracture (frailty) did not find meaningful associations, supporting the robustness of our estimates. Future studies could consider instrumental variable analyses to further address potential selection bias and confounding by indication. Second, the administrative data did not capture information on migraine aura, frequency and severity, which could better quantify the migraine-associated cardiovascular risk (*Supplement Figure 1b*). Although the probabilistic bias analysis accounted for unmeasured confounding, they were based on assumptions and parameters informed by the literature, and should be interpreted with caution. Third, although outpatient encounter diagnoses within VHA demonstrated high sensitivity and specificity to capture cardiovascular comorbidities^39^, false-negative misclassification cannot be eliminated. By restricting the study sample among those with primary care visits and leveraging VA Community Care and CMS claims, the risk of misclassification bias is likely minimal. Lastly, gaps between prescription fills were defined “retrospectively” in this study.^40^ If patients did not redeem a new prescriptions within the 90-day grace period, follow-up was censored at day 0 of the grace period. This approach may introduce a dependency on treatment status on future prescription refills. However, the conclusions of this study are not biased as they were drawn based on intention-to-treat analyses.

In summary, findings from this study suggest that initiation of lisinopril/candesartan may be associated with a higher risk of MACE, particularly MI, with greater risk observed in migraine patients with minimal risk of atherosclerotic diseases. In contrast, aCGRP treatment appears safe regarding cardiovascular events and may reduce the risk of MI. These **temporal** associations are biologically **plausible** given the roles of CGRP and SP in neurogenic inflammation, and **consistent** with prior observational studies comparing aCGRP treatment to neurotoxins. The **strength** of association in **specific** subgroups with minimal cardiovascular indications for ACEI/ARB use, along with **analogous** findings in chronic pain populations and post-fracture ACEI users, further supports this hypothesis. The contrast between aCGRP treatment (which suppresses neurogenic inflammation) and lisinopril (which may aggravate it via reduced degradation of SP) mimics **experimental manipulation**. However, **coherence** with findings from randomized trials and existing clinical guidelines, which are not migraine-specific, remains limited. Future research is needed to assess **dose-response** relationships, replicate these findings across diverse populations with migraine, and further elucidate the role of neurogenic inflammation in migraine-associated non-atherosclerotic cardiovascular risk.

## Data Availability

The data used for this study are available with an approved study protocol by the Department of Veterans Affairs. The data are not publicly available due to regulations and ethics agreements.

## Non-standard Abbreviations and Acronyms

aCGRP: anti-calcitonin gene-related peptide
CDW: Corporate Data Warehouse
ICH/SAH: intracerebral and subarachnoid hemorrhage
IPTaW: inverse probability of treatment adherence weights
IPTiW: inverse probability of treatment initiating weights
MACE: major adverse cardiovascular events
MPR: medication possession ratio
RD: risk difference
SBP: systolic blood pressure
SI: simulation interval
SMD: standardized mean difference
SP: substance P
VA: U.S. Department of Veterans Affairs

## Acknowledgement

This research was supported by the resources and computational environment of the VA Informatics and Computing Infrastructure (VINCI) (DART# 2024-02-057-D), as well as by the use of Centers for Medicare and Medicaid data provided by the VA Information Resource Center (VIReC) (DUI# Wang-05-1). This publication does not necessarily represent the views of the Department of Veterans Affairs or the United States Government.

