## Supplementary for "Migraine, Neurogenic Inflammation, and Cardiovascular Risk: A Pharmacoepidemiologic Study of ACEI/ARB and Anti-CGRP Treatment"

**Supplemental Material**

**Supplement Methods of Probabilistic Bias Analyses for Residual Confounding**

**Supplement Table 1**. Target Trial Specification and Emulation.

**Supplement Table 2**. Distribution of Medications at Initiation and Throughout Follow-Up.

**Supplement Table 3**. Definitions of Migraine, Outcome, and Covariates.

**Supplement Table 4**. Distribution of First Comorbidity Diagnoses Before Baseline and First Outcome Events after Baseline by Data Source.

**Supplement Table 5**. Distribution of Inverse Probability of Treatment Adherence Weights (IPTaW) Estimated Using Baseline and Time-Updated Covariates to Account for Potential Selection Bias Associated with Non-Adherence.

**Supplement Table 6**. Sensitivity Analysis Among 19,250 Lisinopril/Candesartan and 150,096 Topiramate Person-Trials in Migraine Patients Without Cardiovascular Indication between, 10/1/2007 – 9/30/2024.

**Supplement Table 7**. Subgroup Analyses by Age and Systolic Blood Pressure Among Patients Who Initiated Lisinopril/Candesartan or Topiramate, Overall and by Presence of Cardiovascular Indications.

**Supplement Table 8**. Sensitivity Analyses Using Outcome Regressions with Baseline Confounders and Additional Adjustment for Competing Risk of Death in Myocardial Infarction and Ischemic Stroke.

**Supplement Table 9.** Probabilistic Bias Analysis Assessing the Impact of an Unmeasured Confounder.

**Supplement Table 10**. Estimates for Positive and Negative Outcome Controls.

**Supplement Figure 1**. Directed Acyclic Graph Illustrating (a) Potential Confounders in the Association Between Exposure and MACE, and (b) Hypothesized Pathways of Migraine-Associated Cardiovascular Risk via CGRP and Substance P.

**Supplement Figure 2**. Standardized Mean Differences Before and After Applying the Inverse Probability of Treatment Initiation Weights (IPTiW).

**Supplement Figure 3.** Cumulative Incidence Curves of MACE, MI And Ischemic Stroke Among Patients Who Initiated Lisinopril/Candesartan or Topiramate, Stratified by Age And Systolic Blood Pressure.

**Supplement Methods of Probabilistic Bias Analyses for Residual Confounding**

We conducted probabilistic bias analyses^1^ to assess the potential impact of an unmeasured confounder, such as migraine frequency or an unknown cardiovascular risk factor, on the observed associations ${RR}_{obs}$ between migraine preventive treatments (anti-calcitonin gene-related peptide [aCGRP] treatment or lisinopril/candesartan) and the risk of myocardial infraction (MI). Assuming no interaction between the unmeasured confounder and the treatment, the bias-adjusted risk ratio ${RR}_{adj}$ can be estimate at the summary level using the formula:

$${RR}_{adj}={RR}_{obs}\frac{{RR}_{CD}\times p_{0}+(1-p_{0)}}{{RR}_{CD}\times p_{1}+(1-p_{1})}$$

where ${RR}_{CD}$ is the risk ratio associating the unmeasured confounder with the outcome assuming no effect modification by the treatment, and $p_{1}$ and $p_{0}$ are the prevalence of individuals with the confounder in the intervention and topiramate groups respectively. The bias-adjusted ${RR}_{adj}$ were obtained from 50,000 Monte Carlo simulations, and 95% simulation intervals (SIs) were computed to reflect the uncertainty from systematic error and random error.

A series of plausible scenarios of were tested:

1. We considered migraine frequency as a potential unmeasured confounder. We modeled monthly migraine days (MMDs) using negative-binomial distributions with dispersion parameter^2^ $\tau=0.2$, assuming an average of 6 MMDs for patients initiating topiramate^3,4^ $NB(r=5,p=0.455)$, and 10 MMDs for those initiating aCGRP treatment^5,6^ $NB(r=5,p=0.333)$. From these distributions, we calculated the proportion of individuals with ≥ 5 MMDs. The association between migraine frequency and MI is not well established. The Women’s Health Study reported that, compared to women with no history of migraine, those with any history of migraine had an adjusted hazard ratio (HR) of 1.10 (95% CI, 0.70 – 1.73), while those with ≥ weekly attacks had an HR of 1.53 (95% CI, 0.38 – 6.15). The implied contrast between ≥ weekly attacks vs. any migraine history yields an HR of 1.39 (95% CI, 0.32 – 6.01), which was used as the ${RR}_{CD}$.
2. We used data from the Migraine in America Symptoms and Treatment (MAST) cross-sectional study^7^, which reported an odds ratio of 1.63 (95% CI, 1.29 – 2.06) for the association between having 5-9 monthly headache days (MHDs) vs. 1-4 MHDs and the presence of comorbid MI among individuals with migraine. We used this estimate as an alternative value for ${RR}_{CD}$.and assumed same negative-binomial distributions for MHDs.
3. We considered migraine with aura as a potential unmeasured confounder. In the VA population, migraine with aura accounts for less than 20% of patients with a migraine diagnosis based on ICD-9/10 code.^8^ In contrast, randomized trials of aCGRP monoclonal antibodies reported aura in approximately 35 – 47% of participants.^5,9,10^ Accordingly, we specified trapezoidal distributions to model the prevalence of aura: $trapezoid(0.35, 0.40, 0.50, 0.55)$ for the aCGRP group, and $trapezoid(0.20, 0.25, 0.35, 0.40)$ for the topiramate group.^11,12^ A systematic review and meta-analysis^13^ reported an HR of 1.77 (95% CI, 1.37 – 2.28) for individuals with aura and 1.17 (95% CI: 0.99 – 1.39) for those without aura, implying an HR of 1.51 (95% CI 1.12 – 2.04) for the contrast between migraine with aura vs. without aura.
4. We assumed the presence of an unknown cardiovascular risk factor that influences MI risk among patients initiated with lisinopril/candesartan but without a documented cardiovascular indication. Based on estimates from Supplemental Table 8, we considered four levels of ${RR}_{CD}$: 1.40 (1.20 – 1.64), 1.60 (1.37 – 1.87), 2.00 (1.71 – 2.24) and 4.00 (3.42 – 4.68), assuming ${SE}_{\beta}=0.08$. The prevalence of this hypothetical risk factor was randomly drawn from a $uniform(0.7, 1.0)$ distribution for the lisinopril/candesartan group, and from a $uniform(0.1, 0.4)$ distribution for the topiramate group.

**Supplement Table 1. Target Trial Specification and Emulation.**

| **Protocol component** | **Target trial specification** | **Target trial emulation** |
| --- | --- | --- |
| **Eligibility criteria** | - Aged 18 and older - Diagnosed with (incident) migraine disorder - Regular healthcare system contact, defined as having at least one primary care visit in the past 12 months. - No prior use of study medication before migraine diagnosis - No use of study medications or medications in the same class in the past 3 months | In addition, we exclude:   - Prescriptions of galcanezumab (100mg/mL), topiramate combined with phentermine, lisinopril/candesartan combined with hydrochlorothiazide - Use of other ACEIs/ARBs in the lisinopril/candesartan vs. topiramate trial - Use of remigepant, ubrogepant, zavegepant as acute treatment in the topiramate group in aCGRP vs. topiramate trial |
| **Treatment strategies** | - Initiate aCGRP treatment - Initiate lisinopril or candesartan - Initiate topiramate | In addition to the target trial specification:   - Discontinuation defined as a 90-day or longer gap between successive prescriptions |
| **Treatment assignment** | Randomly assigned and aware of assigned strategy at baseline | Randomization is assumed conditional on baseline covariates |
| **Outcome** | Primary outcome was major adverse cardiovascular events; secondary outcomes were MI, ischemic stroke and ICH/SAH, respectively. | Same as for the target trial |
| **Follow-up** | Starts at baseline and ends at the occurrence of MACE, 60 months after the baseline, or administrative end of the study by Sep 30, 2024, whichever happens first. | Same as for the target trial |
| **Causal contrasts** | Intention-to-treat effect and per-protocol effect | Observational analog of intention-to-treat and per-protocol effects |
| **Statistical analysis** | Pooled logistic regression to estimate hazard ratios and standardized risk curves and differences.   - Intention-to-treat analysis: apply treatment initiation weight (IPTiW) to adjust for baseline covariates - Per-protocol analysis: censor patients if and when they deviate from assigned treatment strategy and apply treatment adherence weight (IPTaW) to adjust for baseline and time-updated covariates | Same intention-to-treat and per-protocol analyses |

Abbreviations: ACEIs/ARBs, angiotensin-converting enzyme inhibitors/angiotensin II receptor blockers; aCGRP, anti-calcitonin gene-related peptide; ICH/SAH, intracerebral/subarachnoid hemorrhage; MACE, major adverse cardiovascular events; MI, myocardial infraction.

**Supplement Table 2. Distribution of Medications at Initiation and Throughout Follow-Up.**

| **Lisinopril/Candesartan** | | | **aCGRP** | | |
| --- | --- | --- | --- | --- | --- |
| **Medication** | **At Initiation, n (%)**^a^ | **Entire Follow-up, n (%)** | **Medication** | **At Initiation, n (%)^a^** | **Entire Follow-up, n (%)** |
| Candesartan | 824 (1.7) | 913 (1.9) | Eptinezumab | 35 (0.1) | 84 (0.3) |
| Lisinopril | 47787 (98.3) | 47795 (98.3) | Erenumab | 20503 (80.0) | 20761 (81.0) |
|  |  |  | Fremanezumab | 1348 (5.3) | 2926 (11.4) |
|  |  |  | Galcanezumab | 1681 (6.6) | 2782 (10.9) |
|  |  |  | Atogepant | 639 (2.5) | 1228 (4.8) |
|  |  |  | Rimegepant | 1446 (5.6) | 1820 (7.1) |

Abbreviation: aCGRP, anti-calcitonin gene-related peptide.

^a^ Percentages may not sum to 100% because patients could be prescribed a monoclonal antibody and then switched to a gepant for prevention on the same day at treatment initiation.

**Supplement Table 3. Definitions of Migraine, Outcome, and Covariates.**

|  | **Source** | **Definition** | **Form** | **Code** |
| --- | --- | --- | --- | --- |
| Migraine | CDW | ICD-9/10 codes recorded during at least one outpatient evaluation and management encounter with a physician, nurse practitioner, or physical assistant, or from any hospitalization discharge diagnoses. Evaluation and management encounters were defined using CPT codes (99201-99205, 99211-99215, 99241-99245, 99281-99285, 99385-99387, 99395-99397, 99401-99404, 99371-99373, 99441-99444, 99421-99423). | Binary |  |
| ***Outcomes*** | | | | |
| All-cause mortality | Death Ascertainment file | Date of death from the Master Person Index (MPI) and the Social Security Administration Death Master File (SSA DMF), and VHA healthcare data. | Time-to-event |  |
| MI | CDW, Medicare/Medicaid and Community Care Data. A 30-day gap to reconcile an event from overlapping encounters and claims across data sources. | An inpatient discharge diagnosis recorded in the first, second, or third position after the baseline. | Time-to-event |  |
| Ischemic stroke |  |  |  |  |
| ICH |  |  |  |  |
| SAH |  |  |  |  |
| Influenza | CDW, Medicare/Medicaid and Community Care Data. A 30-day gap to reconcile an event from overlapping encounters and claims across data sources. | An inpatient or outpatient diagnosis, or a positive influenza antigen or PCR test within the VA after baseline. | Time-to-event |  |
| Primary malignancy | CDW, Medicare/Medicaid and Community Care Data | An inpatient or outpatient diagnosis | Time-to-first-event |  |
| Non-pathological fracture | CDW, Medicare/Medicaid and Community Care Data. A 90-day gap to reconcile an event from overlapping encounters and claims across data sources. | An inpatient discharge diagnosis recorded in the first, second, or third position after the baseline. | Time-to-event |  |
| ***Baseline covariates*** | | | | |
| Month of baseline | CDW pharmacy domain | The month when initiating treatment, beginning in Jun, 2018. | Natural spline with 4 knots |  |
| Age | CDW patient domain | Date of baseline subtracted by date of birth. | Natural spline with 4 knots at 35, 45, 60 and 75 years old |  |
| Gender | CDW patient domain | Self-identified gender identity. | Categorical | Men; Women |
| Race | CDW patient domain | Most commonly self-reported race values. | Categorical | American Indian/Alaska Native; Asian; Black; Multi-race; Native Hawaiian/Pacific Islander; White; Unknown |
| Ethnicity | CDW patient domain | Self-reported ethnicity. | Categorical | Hispanics; non-Hispanics |
| Residence | CDW patient domain | Attributed to the geocoded patient location. | Categorical | Urban; non-urban (rural, highly rural, unknown) |
| Area deprivation index | CDW | Attributed to the FIPS code of patient residential location, 2015 version. | Continuous |  |
| Service-connected disability | CDW patient domain | An injury or disease resulting in a disability linked to service in the military. | Binary | Yes/No |
| Smoking status | CDW health factor domain | Health factors mapped to distinct smoking status and further refined to one-per-patient considering most recent smoking health factor in relation to past smoking health factors. | Categorical | Never; current; former; unknown |
| Body mass index | CDW vital status domain | 703 multiplicate the most recent weight in lbs. prior to baseline, then divided by median height across all measures. | Natural spline with 3 knots; and categorical | Underweight; normal; overweight; obesity; unknown |
| Systolic BP | CDW vital status domain | Most recent measures prior to baseline; extreme values capped at the 0.1^st^ percentile. | Natural spline with 2 knots |  |
| Diastolic BP |  |  | Natural spline with 4 knots |  |
| CAN score | CDW | Predicted one-year probability of death | Natural spline with 2 knots |  |
| Primary care visits | CDW outpatient domain | Encounters with primary stop code in 170, 172, 178, 301, 318, 319, 322, 323, 324, 326, 338, 348, 350 and a CPT code in 99201-99205, 99211-99215, 99241-99245, 99281-99285, 99385-99387, 99395-99397, 99401-99404, 99371-99373, 99441-99444, 99421-99423 with a physician, NP or PA in the past 12 months. | Categorical | None; Once; Twice; Three times and more. |
| Flu vaccination | CDW immunization domain | CVX code in 15, 16, 88, 111, 125-128, 135, 140, 141, 144, 149-151, 153, 155, 158, 160, 161, 166, 168, 171, 185, 186, 194, 197, 200-202, 205, 231, 320 in the past 12 months. | Binary | Yes/No |
| Comorbidities | CDW, Medicare/Medicaid and Community Care Data | ICD-9/10 codes recorded during at least two outpatient encounters, or from any hospitalization discharge diagnoses prior to the baseline. | Binary | Yes/No |
| *TBI* | CDW, Medicare/Medicaid, Community Care Data, and CDW health factor domain | A positive TBI screening in addition to ICD-9/10 codes. | Binary | Yes/No |
| Medications | CDW pharmacy domain | Medications released from outpatient pharmacy and were taking at the baseline. | Binary | Yes/No |
| *Neurotoxins* | CDW pharmacy domain and outpatient procedure domain | Medications released from outpatient pharmacy or outpatient CPT codes in 64615, J0585-J0588. | Binary | Yes/No |
| Headache-related ED visits | CDW outpatient domain | Encounters with primary stop code in 130 or 131 associated with a physician, NP or PA in the past 12 months. | Binary | Yes/No |
| Headache-related neurology visits | CDW outpatient domain | Encounters with primary stop code in 315 or 325 associated with a physician, NP or PA in the past 12 months. | Categorical | None; Once; Twice; Three times and more. |
| ***Time-updated covariates*** | | | | |
| Month of follow-up | CDW pharmacy domain | Month of the person-trial since the baseline. | Natural spline with knots at 6, 12, 24, and 48 months |  |
| Body mass index | CDW vital status domain | 703 multiplicate the most recent weight in lbs. prior to the beginning of each person-trial-month, then divided by median height across all measures. | Natural spline with 3 knots |  |
| Systolic BP | CDW vital status domain | Most recent measures prior to each person-trial-month; extreme values capped at the 0.1st percentile. | Natural spline with 2 knots |  |
| Diastolic BP | CDW vital status domain |  | Natural spline with 4 knots |  |
| Any hospitalization | CDW, Medicare/Medicaid and Community Care Data. | Any hospitalization occurred in the past month prior to the beginning of each person-trial-month. | Binary | Yes/No |
| CKD | CDW, Medicare/Medicaid and Community Care Data. | ICD-9/10 codes recorded during at least two outpatient encounters, or from any hospitalization discharge diagnoses prior to the beginning of each person-trial-month. | Binary | Yes/No |
| Heart failure |  |  |  |  |
| Hypertension |  |  |  |  |
| Chronic migraine |  |  |  |  |
| β-blockers | CDW pharmacy domain | Medications released from outpatient pharmacy and were taking at the beginning of each person-trial-month. | Binary | Yes/No |
| Diuretics |  |  |  |  |
| Antiplatelets |  |  |  |  |
| Triptans |  |  |  |  |
| NSAIDs |  |  |  |  |

Abbreviations: BP, blood pressure; CDW, Corporate Data Warehouse; CKD, chronic kidney disease; CPT, Current Procedural Terminology; ED, emergency department; ICD, International Classification of Diseases; ICH, intracerebral hemorrhage; NSAIDs, non-steroidal anti-inflammatory drugs; PCR, polymerase chain reaction; SAH, subarachnoid hemorrhage; TBI, traumatic brain injury; VA, Department of Veterans Affairs.

**Supplement Table 4. Distribution^a^ of First Comorbidy Diagnoses Before Baseline and First Outcome Events after Baseline by Data Source.**

|  | **VA CDW** | **Community Care** | **Medicare** | **Medicaid** | **Total** |
| --- | --- | --- | --- | --- | --- |
| ***Baseline comorbidities*** |  |  |  |  |  |
| Atrial fibrillation/flutter | 3410 (57.2) | 1114 (18.7) | 1414 (23.7) | 131 (2.2) | 5965 |
| Alcohol-related disorder | 35415 (93.4) | 1538 (4.1) | 741 (2.0) | 379 (1.0) | 37931 |
| Cardiomyopathy | 1830 (56.1) | 753 (23.1) | 659 (20.2) | 121 (3.7) | 3260 |
| Chronic inflammatory conditions |  |  |  |  |  |
| Ankylosing spondylitis | 459 (76.8) | 61 (10.2) | 70 (11.7) | 10 (1.7) | 598 |
| HIV | 810 (98.2) | 17 (2.1) | 0 (0.0) | 0 (0.0) | 825 |
| inflammatory bowel disease | 2290 (81.0) | 258 (9.1) | 234 (8.3) | 60 (2.1) | 2826 |
| Psoriatic arthritis | 2196 (84.0) | 205 (7.8) | 176 (6.7) | 41 (1.6) | 2613 |
| Rheumatoid arthritis | 2402 (68.7) | 414 (11.8) | 635 (18.2) | 69 (2.0) | 3495 |
| Systemic lupus erythematosus | 992 (73.8) | 149 (11.1) | 156 (11.6) | 53 (3.9) | 1344 |
| Systemic sclerosis | 83 (66.4) | 19 (15.2) | 22 (17.6) | 2 (1.6) | 125 |
| Chronic kidney disease | 8264 (67.2) | 1569 (12.8) | 2381 (19.4) | 232 (1.9) | 12292 |
| COPD | 12443 (74.8) | 1713 (10.3) | 2302 (13.8) | 331 (2) | 16633 |
| Diabetes | 32952 (86.7) | 1826 (4.8) | 2743 (7.2) | 741 (1.9) | 38001 |
| DVT/PE | 1771 (57.6) | 740 (24.1) | 481 (15.6) | 120 (3.9) | 3074 |
| Dyslipidemia | 95676 (92.6) | 2521 (2.4) | 4614 (4.5) | 941 (0.9) | 103319 |
| Heart failure | 3195 (47.1) | 1754 (25.8) | 1828 (26.9) | 214 (3.2) | 6790 |
| Hypertension | 89809 (94.3) | 2251 (2.4) | 3008 (3.2) | 1039 (1.1) | 95212 |
| Depression | 130385 (95.4) | 3435 (2.5) | 2242 (1.6) | 1010 (0.7) | 136713 |
| Obstructive sleep apnea | 16499 (89.5) | 393 (2.1) | 1219 (6.6) | 334 (1.8) | 18429 |
| Peripheral artery disease | 4889 (53.1) | 1475 (16.0) | 2748 (29.8) | 176 (1.9) | 9211 |
| Substance-related disorder | 22004 (83.4) | 2223 (8.4) | 1751 (6.6) | 554 (2.1) | 26378 |
| Traumatic brain injury ^b^ | 36635 (92.1) | 1982 (5.0) | 1117 (2.8) | 273 (0.7) | 39798 |
| Valvular heart disease | 3160 (43.9) | 2110 (29.3) | 1803 (25.0) | 216 (3.0) | 7203 |
| Chronic migraine | 32849 (82.6) | 5235 (13.2) | 1733 (4.4) | 401 (1.0) | 39751 |
| History of |  |  |  |  |  |
| Myocardial infraction | 1294 (31.6) | 1876 (45.8) | 927 (22.6) | 162 (4.0) | 4096 |
| Ischemic Stroke | 3491 (56.5) | 1633 (26.4) | 985 (16.0) | 160 (2.6) | 6174 |
| Intracerebral hemorrhage | 200 (39.5) | 213 (42.1) | 92 (18.2) | 12 (2.4) | 506 |
| Subarachnoid hemorrhage | 107 (28.2) | 197 (52.0) | 66 (17.4) | 16 (4.2) | 379 |
| ***Outcome events*** |  |  |  |  |  |
| Myocardial infraction | 697 (26.2) | 1661 (62.5) | 335 (12.6) | 42 (1.6) | 2658 |
| Ischemic Stroke | 439 (19.8) | 1618 (72.8) | 187 (8.4) | 23 (1.0) | 2222 |
| Intracerebral hemorrhage | 28 (10.9) | 204 (79.7) | 28 (10.9) | 3 (1.2) | 256 |
| Subarachnoid hemorrhage | 13 (8.1) | 140 (87.0) | 5 (3.1) | 3 (1.9) | 161 |
| Any malignancy | 9319 (76.3) | 2083 (17.1) | 768 (6.3) | 103 (0.8) | 12214 |
| Influenza | 3195 (43.5) | 3615 (49.2) | 458 (6.2) | 166 (2.3) | 7353 |
| Non-pathological fracture | 7707 (47.7) | 7082 (43.9) | 1507 (9.3) | 280 (1.7) | 16142 |

Abbreviations: DVT/PE, deep vein thrombosis/pulmonary embolism.

^a^ Row percentages may exceed 100% due to same-day diagnoses from different data sources.

^b^ The distribution of TBI is based solely on ICD-9/10 codes, without incorporating TBI screening data from the health factor domain.

**Supplement Table 5. Distribution of Inverse Probability of Treatment Adherence Weights (IPTaW) Estimated Using Baseline and Time-Updated Covariates to Account for Potential Selection Bias Associated with Non-Adherence.**

|  | **Min** | **1%** | **5%** | **25%** | **50%** | **75%** | **95%** | **99%** | **Max** |
| --- | --- | --- | --- | --- | --- | --- | --- | --- | --- |
| **Lisinopril/Candesartan** |  |  |  |  |  |  |  |  |  |
| MACE | 0.027800 | 0.487547 | 0.755431 | 0.967021 | 1.000000 | 1.044109 | 1.238241 | 1.559445 | 26.746986 |
| MI | 0.028718 | 0.487940 | 0.754651 | 0.966587 | 1.000000 | 1.044398 | 1.239289 | 1.561759 | 44.343155 |
| Ischemic Stroke | 0.028983 | 0.489070 | 0.755180 | 0.966589 | 1.000000 | 1.044321 | 1.238660 | 1.559415 | 45.985806 |
| ICH/SAH | 0.029490 | 0.488801 | 0.754678 | 0.966453 | 1.000000 | 1.044503 | 1.239754 | 1.562415 | 49.967112 |
| **aCGRP** |  |  |  |  |  |  |  |  |  |
| MACE | 0.020513 | 0.480639 | 0.766852 | 0.975143 | 1.001111 | 1.043392 | 1.220741 | 1.518931 | 42.704187 |
| MI | 0.020730 | 0.480676 | 0.765874 | 0.974907 | 1.001027 | 1.043590 | 1.221840 | 1.521746 | 38.669784 |
| Ischemic Stroke | 0.021421 | 0.482504 | 0.766456 | 0.974906 | 1.001034 | 1.043572 | 1.221190 | 1.518585 | 36.695729 |
| ICH/SAH | 0.021319 | 0.481533 | 0.765609 | 0.974835 | 1.001055 | 1.043724 | 1.222233 | 1.521865 | 38.331381 |

Abbreviations: aCGRP, anti-calcitonin gene-related peptide; ICH/SAH, intracerebral/subarachnoid hemorrhage; MACE, major adverse cardiovascular events; MI, myocardial infraction.

**Supplement Table 6. Sensitivity Analysis Among 19,250 Lisinopril/Candesartan and 150,096 Topiramate Person-Trials in Migraine Patients Without Cardiovascular Indication ^a^ between, 10/1/2007 – 9/30/2024.**

|  | **5-year cumulative incidence** | | **Risk differences (95%CI)** | **Risk ratio (95% CI)** | **Overall HR (95% CI) ^b^** |
| --- | --- | --- | --- | --- | --- |
|  | **Intervention, % (95% CI)** | **Topiramate, % (95% CI)** |  |  |  |
| **MACE** | 3.74 (3.17, 4.37) | 2.20 (2.08, 2.31) | 1.54 (0.95, 2.16) | 1.70 (1.43, 1.98) | 1.70 (1.44, 2.03) |
| MI | 1.43 (1.02, 1.94) | 0.51 (0.45, 0.58) | 0.92 (0.52, 1.43) | 2.80 (2.02, 3.89) | 2.75 (2.01, 3.77) |
| Ischemic stroke | 0.56 (0.36, 0.84) | 0.49 (0.43, 0.54) | 0.07 (-0.14, 0.36) | 1.15 (0.73, 1.72) | 1.24 (0.76, 2.02) |
| ICH/SAH | 0.14 (0.08, 0.12) | 0.10 (0.07, 0.24) | 0.04 (-0.04, 0.14) | 1.40 (0.63, 2.44) | 1.27 (0.69, 2.35) |

Abbreviations: CI, confidence interval; HR, hazard ratio; ICH/SAH, intracerebral/subarachnoid hemorrhage; MACE, major adverse cardiovascular events; MI, myocardial infraction.

^a^ Patients with hypertension, history of myocardial infraction, heart failure or chronic kidney disease were excluded.

^b^ Robust sandwich estimator.

**Supplement Table 7. Subgroup Analyses by Age and Systolic Blood Pressure Among Patients Who Initiated Lisinopril/Candesartan or Topiramate, Overall and by Presence of Cardiovascular Indications.**

|  | **Overall** | | | **Without cardiovascular indications** | | |
| --- | --- | --- | --- | --- | --- | --- |
|  | **MACE**  HR (95% CI) | **MI**  HR (95% CI) | **Ischemic Stroke**  HR (95% CI) | **MACE**  HR (95% CI) | **MI**  HR (95% CI) | **Ischemic Stroke**  HR (95% CI) |
| **Age, y** |  |  |  |  |  |  |
| <40 | 1.63 (1.16, 2.29) | 3.18 (1.63, 6.20) | 1.51 (0.63, 3.63) | 1.77 (1.16, 2.69) | 3.85 (1.59, 9.34) | 1.53 (0.32, 7.44) |
| 40-54 | 1.09 (0.89, 1.33) | 1.12 (0.74, 1.70) | 0.77 (0.54, 1.10) | 1.65 (1.27, 2.14) | 2.73 (1.74, 4.28) | 0.96 (0.61, 1.50) |
| 55+ | 1.06 (0.94, 1.20) | 1.04 (0.81, 1.34) | 1.00 (0.79, 1.27) | 1.56 (1.28, 1.90) | 1.98 (1.39, 2.83) | 1.23 (0.82, 1.86) |
| **SBP, mmHg** |  |  |  |  |  |  |
| <130 | 1.63 (1.43, 1.86) | 1.86 (1.44, 2.39) | 1.56 (1.15, 2.12) | 2.00 (1.56, 2.56) | 4.02 (2.60, 6.23) | 1.54 (0.78, 3.02) |
| 130-139 | 1.16 (1.00, 1.34) | 1.46 (1.11, 1.92) | 0.94 (0.71, 1.23) | 1.38 (1.08, 1.77) | 2.04 (1.29, 3.23) | 0.91 (0.55, 1.50) |
| 140+ | 0.79 (0.65, 0.97) | 0.73 (0.49, 1.09) | 0.64 (0.47, 0.88) | 1.19 (0.91, 1.56) | 1.23 (0.75, 2.03) | 0.74 (0.46, 1.20) |

Abbreviations: CI, confidence interval; HR, hazard ratio; MACE, major adverse cardiovascular events; MI, myocardial infraction; SBP, systolic blood pressure.

**Supplement Table 8. Sensitivity Analyses Using Outcome Regressions with Baseline Confounders and Additional Adjustment for Competing Risk of Death in Myocardial Infarction and Ischemic Stroke.**

|  | **Lisinopril/Candesartan** | | | **aCGRP** | | |
| --- | --- | --- | --- | --- | --- | --- |
|  | **MACE**  HR (95% CI) | **MI**  HR (95% CI) | **Ischemic Stroke**  HR (95% CI) | **MACE**  HR (95% CI) | **MI**  HR (95% CI) | **Ischemic Stroke**  HR (95% CI) |
| Intervention vs. topiramate | 1.25 (1.17, 1.34) | 1.48 (1.31, 1.67) | 1.12 (0.98, 1.28) | 1.00 (0.91, 1.09) | 0.82 (0.70, 0.97) | 0.96 (0.81, 1.15) |
| Gender, women vs. men | 0.85 (0.78, 0.92) | 0.75 (0.65, 0.86) | 1.21 (1.06, 1.38) | 0.84 (0.77, 0.91) | 0.75 (0.64, 0.88) | 1.21 (1.05, 1.39) |
| Smoking, Current vs. never | 1.43 (1.34, 1.53) | 1.60 (1.42, 1.81) | 1.37 (1.19, 1.56) | 1.42 (1.32, 1.53) | 1.60 (1.40, 1.82) | 1.45 (1.27, 1.67) |
| Alcohol-related disorder | 1.12 (1.05, 1.20) | 0.96 (0.85, 1.09) | 0.94 (0.82, 1.07) | 1.09 (1.01, 1.17) | 0.92 (0.81, 1.06) | 0.98 (0.85, 1.12) |
| Atrial fibrillation/flutter | 1.04 (0.94, 1.16) | 1.00 (0.83, 1.20) | 1.23 (1.01, 1.51) | 1.11 (0.99, 1.17) | 0.96 (0.78, 1.17) | 1.14 (0.91, 1.42) |
| Chronic kidney disease | 1.11 (1.02, 1.21) | 1.28 (1.11, 1.47) | 0.99 (0.83, 1.17) | 1.25 (1.15, 1.35) | 1.47 (1.29, 1.69) | 1.23 (1.05, 1.43) |
| Diabetes | 1.24 (1.17, 1.33) | 1.12 (1.00, 1.25) | 1.45 (1.28, 1.64) | 1.27 (1.18, 1.37) | 1.16 (1.02, 1.32) | 1.56 (1.36, 1.78) |
| Dyslipidemia | 1.12 (1.04, 1.20) | 1.52 (1.33, 1.74) | 1.05 (0.91, 1.21) | 1.09 (1.00, 1.18) | 1.46 (1.24, 1.70) | 1.06 (0.91, 1.24) |
| Hypertension | 1.35 (1.25, 1.46) | 1.43 (1.24, 1.66) | 1.65 (1.41, 1.92) | 1.60 (1.46, 1.74) | 2.00 (1.67, 2.40) | 1.84 (1.56, 2.16) |
| Obstructive sleep apnea | 1.00 (0.92, 1.08) | 1.14 (1.00, 1.30) | 1.04 (0.89, 1.21) | 1.00 (0.92, 1.08) | 1.01 (0.88, 1.16) | 0.87 (0.74, 1.01) |
| Substance-related disorder | 1.34 (1.25, 1.44) | 1.24 (1.10, 1.41) | 1.27 (1.10, 1.47) | 1.33 (1.24, 1.44) | 1.08 (0.94, 1.23) | 1.26 (1.09, 1.46) |
| History of MI | 2.00 (1.91, 2.20) | 4.44 (3.83, 5.14) | 1.28 (1.04, 1.57) | 1.82 (1.63, 2.03) | 3.41 (2.89, 4.03) | 1.20 (0.96, 1.50) |
| History of ischemic stroke | 1.79 (1.64, 1.96) | 1.37 (1.17, 1.61) | 3.86 (3.31, 4.50) | 1.80 (1.64, 1.97) | 1.22 (1.03, 1.45) | 3.76 (3.23, 4.38) |
| Triptans | 0.88 (0.80, 0.98) | 0.92 (0.77, 1.10) | 0.77 (0.62, 0.94) | 0.83 (0.76, 0.91) | 0.87 (0.74, 1.02) | 0.78 (0.65, 0.92) |
| NSAIDs | 1.02 (0.94, 1.11) | 0.95 (0.82, 1.09) | 0.91 (0.78, 1.07) | 1.03 (0.95, 1.12) | 0.95 (0.82, 1.11) | 0.89 (0.76, 1.04) |
| E-value | 1.81 (1.62, UCL) | 2.32 (1.95, UCL) | 1.49 (1.00, UCL) | NA | 1.74 (1.21, UCL) | 1.25 (1.00, UCL) |

Multivariable pooled logistic regression models adjusted for trial number, age, gender, race, ethnicity, residency, smoking status, BMI, SBP, DBP, CAN score, receiving influenza vaccination; diagnoses of alcohol-related disorder (ARD), atrial fibrillation, cardiomyopathy, chronic kidney disease, chronic obstructive pulmonary disease, diabetes, dyslipidemia, heart failure, hypertension, depression, obstructive sleep apnea, peripheral arterial disease, substance-related disorder, traumatic brain injury, valvular heart disease, chronic migraine; history of myocardial infraction (MI), ischemic stroke, intracerebral /subarachnoid hemorrhage; use of β-blockers, calcium channel blockers, diuretics or other antihypertensives, anticoagulants, antiplatelets, lipid-modification agents, hormonal replace therapy (HRT), SGLT-2 inhibitors, antipsychotics, triptans, non-steroid antiinflammation drugs (NSAIDs), other anticonvulsants, tricyclic antidepressants, or neurotoxins; migraine duration, headache-related emergency room or neurology visit; use of any aCGRP treatment in the lisinopril/candesartan vs. topiramate group, and use of any ACEIs/ARBs in the aCGRP vs. topiramate group. Standardized errors were adjusted using robust sandwich estimator. Competing risk from death were accounted by estimating the sub-distribution hazards.

The use of lisinopril/candesartan was associated with 48% higher risk of MI (HR 1.48; 95% CI 1.31-1.67). The e-value for this was 2.32 with a lower confidence interval limit of 1.95, meaning that residual confounding could explain the observed association if there exists an unmeasured confounding variable having a relative risk association at least 2.72 with use of lisinopril/candesartan and with MI. The E-value for the lower limit of the confidence interval was 1.95. Given that this risk ratio was much greater than known MI risk factors examined in the model, such as currently smoking, CKD, diabetes, hypertension, or dyslipidemia, it is unlikely that an unmeasured confounder exists that could overcome the association of lisinopril/candesartan use observed in this analysis.

Abbreviation: UCL, upper confidence level.

**Supplement Table 9.** Probabilistic Bias Analysis Assessing the Impact of an Unmeasured Confounder.

| Sample | Treatment | Outcome | ${RR}_{obs}$ (95% CI) | UBC | $p_{1}$ Distribution | $p_{0}$ Distribution | ${RR}_{CD}$ (95% CI) | ${RR}_{adj}$ (95% SI) |
| --- | --- | --- | --- | --- | --- | --- | --- | --- |
| Overall | aCGRP | MI | 0.80 (0.62, 1.02) | MMDs ≥ 5 | $NB(5,0.333)$ | $NB(5,0.455)$ | 1.39 (0.32, 6.01) | 0.88 (0.62, 1.19) |
| Overall | aCGRP | MI | 0.80 (0.62, 1.02) | MHDs ≥ 5 | $NB(5,0.333)$ | $NB(5,0.455)$ | 1.63 (1.29, 2.06) | 0.92 (0.71, 1.18) |
| Overall | aCGRP | MI | 0.80 (0.62, 1.02) | Aura | $TPZ(0.35,0.40,$  $0.50,0.55)$ | $TPZ(0.20,0.25,$  $0.35,0.40)$ | 1.51 (1.12, 2.04) | 0.90 (0.69, 1.16) |
| Without documented CV indications | Lisinopril Candesartan | MI | 2.75 (2.01, 3.77) | CV risk factor | $U(0.7, 1.0)$ | $U(0.1, 0.4)$ | 1.40 (1.20, 1.64) | 2.29 (1.61, 3.14) |
| Without documented CV indications | Lisinopril Candesartan | MI | 2.75 (2.01, 3.77) | CV risk factor | $U(0.7, 1.0)$ | $U(0.1, 0.4)$ | 1.60 (1.37, 1.87) | 2.13 (1.49, 2.93) |
| Without documented CV indications | Lisinopril Candesartan | MI | 2.75 (2.01, 3.77) | CV risk factor | $U(0.7, 1.0)$ | $U(0.1, 0.4)$ | 2.00 (1.71, 2.24) | 1.90 (1.31, 2.65) |
| Without documented CV indications | Lisinopril Candesartan | MI | 2.75 (2.01, 3.77) | CV risk factor | $U(0.7, 1.0)$ | $U(0.1, 0.4)$ | 4.00 (3.42, 4.68) | 1.45 (0.90, 2.14) |

Abbreviations: aCGRP, anti-calcitonin gene-related peptide; CI, confidence interval; CV, cardiovascular; MI, myocardial infraction; MHDs, monthly headache days; MMDs, monthly migraine days; NB, negative-binomial; RR, risk ratio; TPZ, trapezoid; U, uniform; UBC, unmeasured binary confounder.

**Supplement Table 10. Estimates for Positive and Negative Outcome Controls.**

| **Time-to-event outcomes** | **5-year cumulative incidence** | | **Risk differences**  **(95% CI)** | **Risk ratio (95% CI)** | **Overall HR (95% CI) ^a^** |
| --- | --- | --- | --- | --- | --- |
|  | **Intervention, %**  **(95% CI)** | **Topiramate, %**  **(95% CI)** |  |  |  |
| **Lisinopril/Candesartan** |  |  |  |  |  |
| Influenza | 5.97 (5.33, 6.66) | 5.61 (5.38, 5.85) | 0.36 (-0.37, 1.14) | 1.06 (0.94, 1.20) | 1.08 (0.96, 1.23) |
| Non-pathological fracture | 12.13 (11.39, 12.98) | 11.49 (11.09, 12.01) | 0.63 (-0.27, 1.66) | 1.06 (0.98, 1.15) | 1.06 (0.98, 1.14) |
| Primary malignancy | 4.68 (4.16, 5.16) | 4.02 (3.75, 4.34) | 0.66 (0.05, 1.24) | 1.16 (1.01, 1.32) | 1.24 (1.09, 1.42) |
| **aCGRP** |  |  |  |  |  |
| Influenza | 5.76 (4.79, 6.87) | 5.87(5,68, 6.05) | -0.10 (-1.12, 0.97) | 0.98 (0.81, 1.16) | 0.99 (0.87, 1.13) |
| Non-pathological fracture | 12.24 (10.93, 13.83) | 11.20 (10.95, 11.44) | 1.04 (-0.21, 2.61) | 1.09 (0.98, 1.23) | 1.13 (1.01, 1.26) |
| Primary malignancy | 4.62 (3.69, 5.76) | 3.71 (3.56, 3.87) | 0.91 (-0.03, 2.07) | 1.25 (0.99, 1.56) | 1.32 (1.07, 1.63) |
| **Continuous outcome** ^b^ | **1-year estimate** | | **Differences**  **(95% CI)** |  |  |
|  | **Intervention**  **(95% CI)** | **Topiramate**  **(95% CI)** |  |  |  |
| **Lisinopril/Candesartan** |  |  |  |  |  |
| Weight change, kg | -0.39 (-0.47, -0.31) | -1.29 (-1.35, -1.23) | 0.90 (0.80, 0.99) |  |  |
| **aCGRP** |  |  |  |  |  |
| Weight change, kg | -0.70 (-0.79, -0.60) | -1.49 (-1.54, -1.43) | 0.79 (0.68, 0.90) |  |  |

Abbreviations: aCGRP, anti-calcitonin gene-related peptide; CI, confidence interval; HR, hazard ratio.

^a^ Robust sandwich estimator

^b^ Weighted Mixed-effects models

**Supplement Figure 1.** **Directed Acyclic Graph Illustrating (a) Potential Confounders in the Association Between Exposure and MACE, and (b) Hypothesized Pathways of Migraine-Associated Cardiovascular Risk via CGRP and Substance P.**

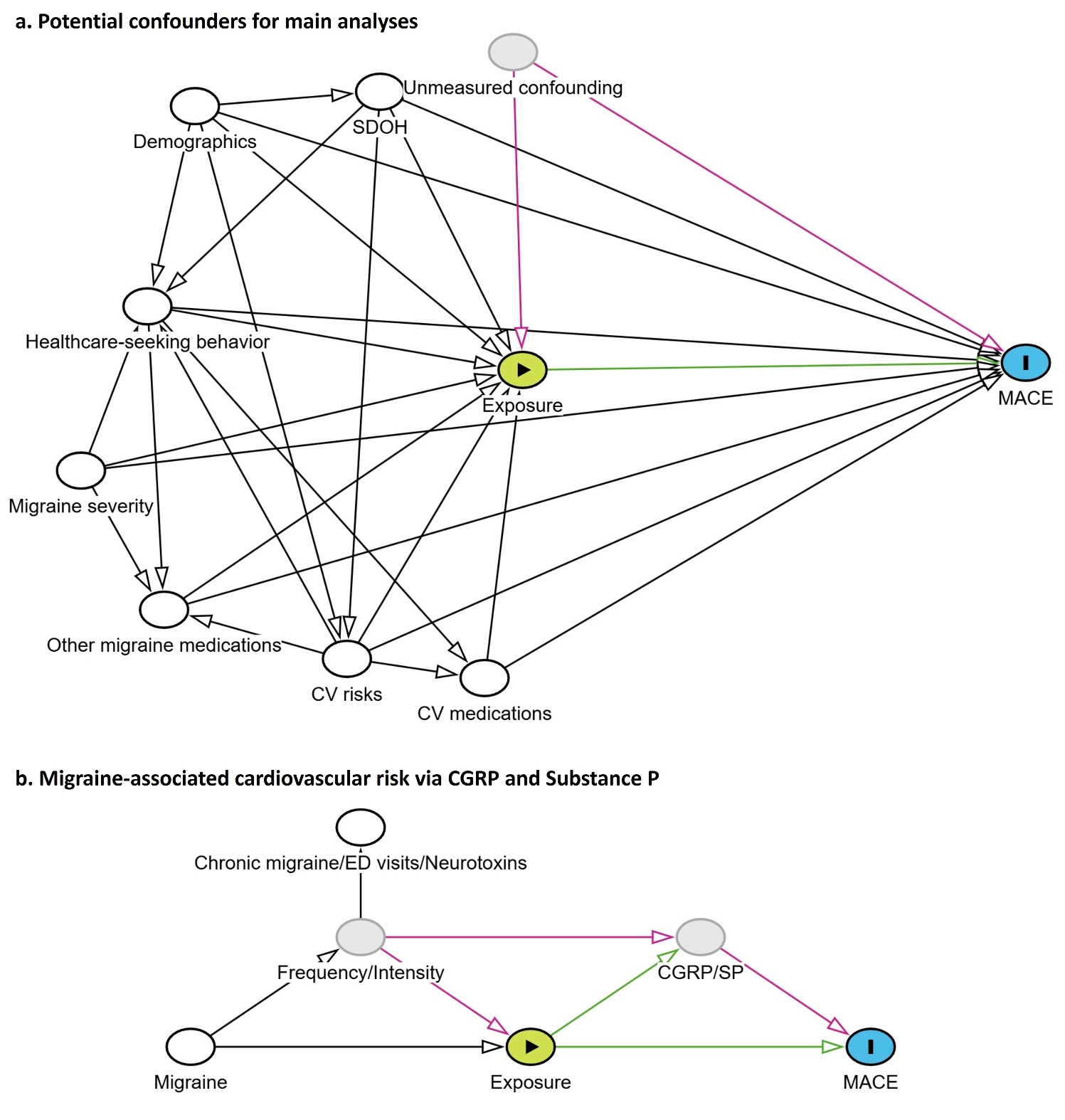

White nodes represent confounding variables that has been adjusted for; the grey node represents unmeasured confounding variable(s). Green arrow indicates causal path; purple arrow illustrates bias path; black arrows indicate paths that have been blocked by adjustment.

Demographics include age, gender, race and ethnicity. Social determinants of health (SDOH) include residency. Healthcare-seeking behavior is proxied by receipt of influenza vaccination, primary care encounters, and Care Assessment Need (CAN) score. Migraine severity is measured by the migraine duration, diagnosis of chronic migraine, headache-related emergency room and neurology visits, and treatment with neurotoxins. Other migraine medications include both abortive (e.g., triptans, NSAIDs, zavegepant) and preventive treatments (e.g., valproates, tricyclic antidepressants). Cardiovascular risk factors include smoking status, BMI, blood pressure, cardiovascular comorbidities (e.g., diabetes, dyslipidemia, CKD), and history of MACE. Cardiovascular-related medications such as antiplatelets, β-blockers, lipid modifying agents.

**Supplement Figure 2. Standardized Mean Differences Before and After Applying the Inverse Probability of Treatment Initiation Weights (IPTiW).**

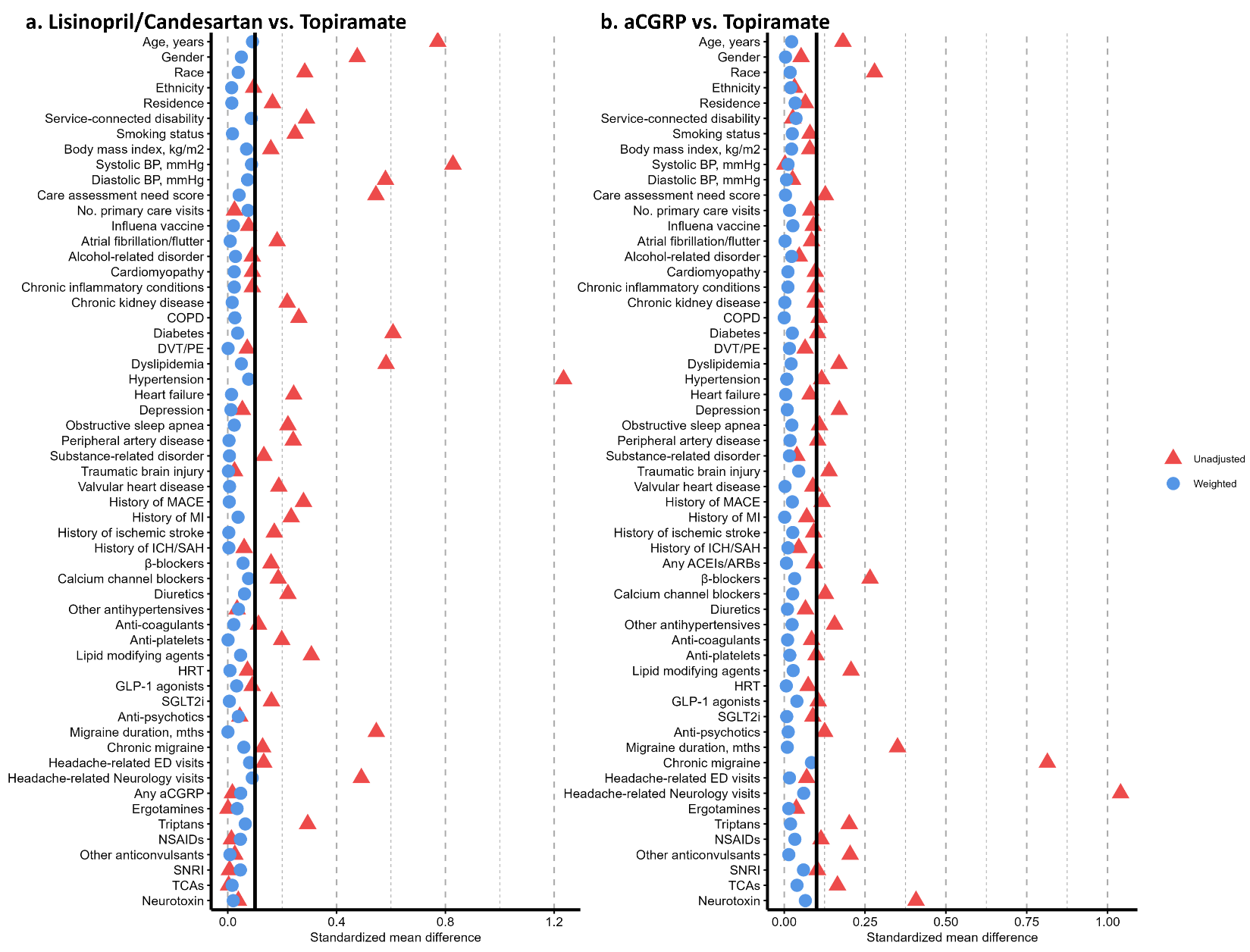

Logistic regression models included trial number, age, gender, race, ethnicity, residency, smoking status, BMI, SBP, DBP, CAN score, receiving influenza vaccination; diagnoses of alcohol-related disorder (ARD), atrial fibrillation, cardiomyopathy, chronic kidney disease (CKD), chronic obstructive pulmonary disease, diabetes, dyslipidemia, heart failure, hypertension, depression, obstructive sleep apnea, peripheral arterial disease, substance-related disorder (SRD), traumatic brain injury, valvular heart disease, chronic migraine; history of myocardial infraction (MI), ischemic stroke, intracerebral /subarachnoid hemorrhage; use of β-blockers, calcium channel blockers, diuretics or other antihypertensives, anticoagulants, antiplatelets, lipid-modification agents, hormonal replace therapy (HRT), SGLT-2 inhibitors, antipsychotics, triptans, non-steroid antiinflammation drugs (NSAIDs), other anticonvulsants, tricyclic antidepressants, or neurotoxins; migraine duration, headache-related emergency room or neurology visit; use of any aCGRP treatment in the lisinopril/candesartan vs. topiramate group, and use of any ACEIs/ARBs in the aCGRP vs. topiramate group.

**Supplement Figure 3. Cumulative Incidence Curves of MACE, MI And Ischemic Stroke Among Patients Who Initiated Lisinopril/Candesartan or Topiramate, Stratified by Age And Systolic Blood Pressure.**

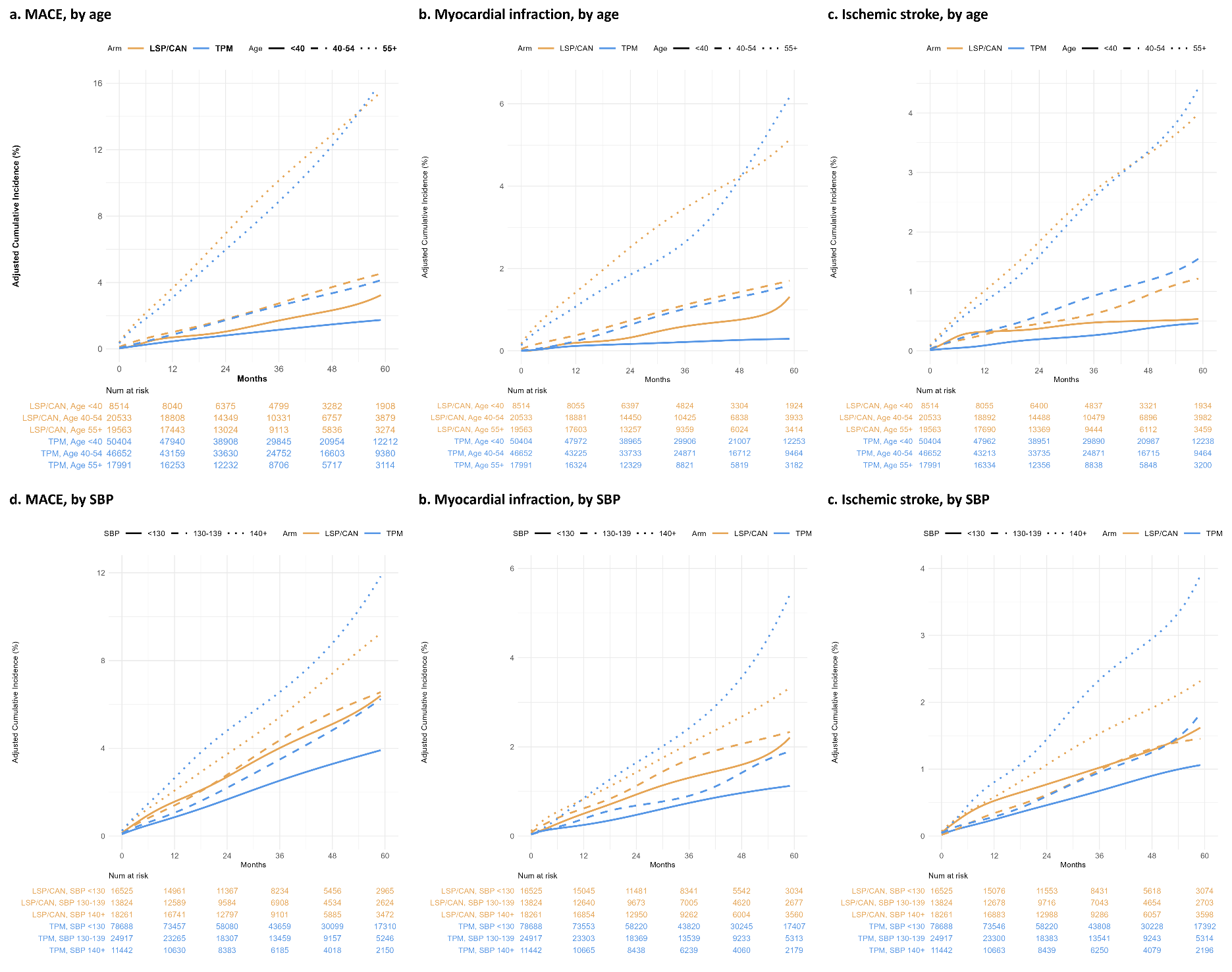
